# Plasma phosphorylated tau species across neurodegenerative and cerebrovascular diseases: An ONDRI study

**DOI:** 10.64898/2026.11.06.26367445

**Authors:** Erlan Sanchez, Paul Connon, Maurice Pasternak, Agessandro Abrahao, Julia Keith, Allison A. Dilliott, Andrée-Ann Baril, Andrew Frank, Anthony E. Lang, Bruce G. Pollock, Connie Marras, Corinne E. Fischer, Dallas Seitz, Dar Dowlatshahi, David A. Grimes, David F. Tang-Wai, Demetrios J. Sahlas, Ekaterina Rogaeva, Elizabeth Finger, Kübra Tan, Malcolm A. Binns, Maria C. Tartaglia, Michael Borrie, Morris Freedman, Rafaella A. Gonçalves, Richard H. Swartz, Robert A. Hegele, Roger A. Dixon, Saira Saeed Mirza, Sali M.K. Farhan, Sandra E. Black, Sanjeev Kumar, Stephen H. Pasternak, Tarek K. Rajji, Thomas Steeves, Walter Swardfager, Henrik Zetterberg, Hlin Kvartsberg, Douglas P. Munoz, Lorne Zinman, Michael J. Strong, Mario Masellis, the Ontario Neurodegenerative Disease Research Initiative (ONDRI)

## Abstract

Elevated plasma phosphorylated tau (p-tau) species are proposed as specific biomarkers of Alzheimer’s disease (AD) pathology but have also been reported in amyotrophic lateral sclerosis (ALS). Their profile across neurodegenerative and cerebrovascular diseases, as well as their discriminatory value when combined with other plasma biomarkers, remain uncertain.

We studied 551 participants (ages 40.1 to 87.1 years; 207 women), comprising 506 patients from the Ontario Neurodegenerative Disease Research Initiative (ONDRI) multi-site cohort and 45 amyloid-β (Aβ) PET-negative cognitively unimpaired controls from the Brain-Eye Amyloid Memory study. Ten diagnostic groups were studied: mild cognitive impairment (MCI), AD, Parkinson’s disease subdivided into cognitively unimpaired (CU), MCI, and dementia, frontotemporal dementia (FTD), cerebrovascular disease (CVD) subdivided into cognitively unimpaired (CU) and cognitively impaired (CI), ALS, and the control group. Baseline visit plasma measurements included p-tau217, p-tau181, Aβ42/40, neurofilament light chain (NfL), and glial fibrillary acidic protein (GFAP). Cross-sectional analyses compared biomarker levels as well as p-tau217-based inference of amyloid positivity status across diagnostic groups. Receiver operating characteristic-area under the curve (ROC-AUC) analyses ascertained the discriminatory capacity of these biomarkers and ratio combinations. Within the ALS group, baseline biomarker measures were also tested for association against baseline clinical features as well as longitudinal measures of neuropsychological change.

P-tau217, p-tau181, GFAP, and NfL differed across groups (all P < 0.001). Compared to controls, p-tau217 was elevated in AD and ALS, p-tau181 and GFAP only in AD, and NfL in ALS and FTD. Inferred amyloid-β positivity was more probable in MCI, AD, ALS, and cognitively impaired CVD groups compared to controls (all P < 0.05). The p-tau217 to NfL ratio discriminated ALS from AD with an ROC-AUC of 0.92 (95% CI 0.85-0.97) compared to p-tau217 alone at 0.65 (95% CI 0.52-0.76). No ALS clinical or longitudinal association survived correction for multiple comparisons.

Elevated p-tau217 and inferred amyloid-β positivity in ALS provide further evidence that the assay signal is not necessarily specific for AD pathology. However, the use of p-tau217 in combination with NfL provides improved discriminatory power between ALS and AD, supporting the use of plasma biomarker combinations rather than sole reliance on individual plasma biomarkers. Inferred amyloid-β positivity in cognitively impaired CVD may indicate AD and/or cerebral amyloid angiopathy co-pathology. Currently available plasma biomarkers should therefore be used as diagnostic complements and interpreted within clinical context rather than as standalone molecular diagnoses.

## Introduction

Blood-based biomarkers of Alzheimer’s disease (AD) pathology are poised to transform not only how AD is diagnosed and managed but also related neurodegenerative diseases, which often harbour amyloid-related AD co-pathology, allowing clinicians access to a less-invasive, lower-cost method of objective diagnosis than cerebrospinal fluid (CSF) sampling or positron emission tomography (PET) alone. This is increasingly important as the field moves toward biological, molecularly defined classifications of AD,*^1^* as well as Parkinson’s disease (PD), with the latter using promising emerging seed amplification assays specific for α-synuclein pathology in CSF or skin samples.*^2,3^* Moreover, as disease-modifying therapies enter clinical use,*^4,5^* identification of early-stage disease and potential neurodegenerative comorbidities is crucial for timely and appropriate instigation of treatment, often before a confident clinical diagnosis is possible.

In contrast to CSF, plasma β-amyloid (Aβ) biomarker assays exhibit only modest discriminative ability due to the relatively narrow dynamic range separating amyloid-positive from amyloid-negative individuals.*^6,7^* By comparison, plasma p-tau217 and p-tau181 demonstrate much larger group separation and consequently have emerged as leading blood-based biomarker candidates for AD.*^8,9^* These species represent an early physiological reaction to Aβ plaques, rather than directly reflecting neurofibrillary tangles, and thus are best understood as surrogate markers of brain amyloid status.*^10^* Furthermore, plasma p-tau217 and p-tau181 assays have shown promising utility in distinguishing AD from other neurodegenerative disorders. This has led proponents to speculate on their potential application as screening tools for AD irrespective of disease stage.*^11,12^* Nonetheless, autopsy studies,*^13^* in addition to studies investigating CSF AD biomarkers and amyloid PET, have shown that Aβ pathology is also evident in Lewy body disorders,*^14^* vascular cognitive impairment,*^15^* cerebral amyloid angiopathy,*^16^* frontotemporal dementia and atypical parkinsonian syndromes,*^17^* and limbic-predominant age-related TDP-43 encephalopathy (LATE).*^18^* The extent of overlap in plasma p-tau distribution across neurodegenerative disorders beyond AD continues to be elucidated,*^11,17,19–26^* including by our group,*^27^* although the precise clinical implications of such co-pathology are yet to be fully characterised.

A cautionary note regarding the interpretation of blood-based biomarkers is that they remain an indirect measure of pathology and are sampled in a fluid remote from the central nervous system (CNS). As such, an elevated concentration does not inherently establish where the underlying protein originates. This limitation has become relevant for p-tau, because plasma p-tau181 and p-tau217 are now reported to be elevated in amyotrophic lateral sclerosis (ALS) by several independent groups.*^28–30^* The finding is unexpected, since ALS is generally characterised by TDP-43 rather than amyloid and tau pathology and CSF p-tau is consistently normal,*^31^* which suggests that part of the plasma signal in this setting may arise peripherally and confound measurement of the CNS-derived fraction. It is also recognized that tau functions extend beyond microtubule stabilization to include RNA binding and association with stress granules,*^32,33^* but whether these contribute to the p-tau plasma signal in ALS remains to be determined.

There are additional blood-based biomarkers that have gained clinical interest. Neurofilament light chain (NfL) is a non-specific marker of neuroaxonal injury that reflects neurodegeneration across many neurological conditions,*^34^* and glial fibrillary acidic protein (GFAP) reflects astrocytic reactivity and contributes to disease staging in AD.*^35,36^* Both are included as non-core markers of neurodegeneration and neuroinflammation, respectively, in the revised NIA-AA framework.*^1^*

The behaviour of these biomarkers across the full neurodegenerative and cerebrovascular disease spectrum, and their ability to discriminate between clinically diagnosed groups requires further investigation. We therefore measured plasma p-tau217, p-tau181, Aβ42, Aβ40, NfL, and GFAP across patients spanning mild cognitive impairment (MCI), AD dementia, PD classified as cognitively unimpaired (PD-CU), PD with mild cognitive impairment (PD-MCI) or PD with dementia (PD-D), frontotemporal dementia (FTD) and related disorders, cerebrovascular disease (CVD) classified as cognitively unimpaired (CVD-CU) or with cognitive impairment (CVD-I), and ALS, together with amyloid-β PET-negative cognitively unimpaired controls (HC). These analytes were compared between these ten clinically defined diagnostic groups through both statistical group-difference analyses and receiver operating characteristic-area under the curve (ROC-AUC) diagnostics. To assess possible amyloid co-pathology, we also inferred Aβ positivity from plasma p-tau217 using the two-step thresholding approach of Brum et al.,*^37^* as applied to the ALZpath assay by Ashton et al.,*^12^* and examined this across all clinical diagnostic groups.

## Methods

### Study sample and recruitment

This study included patients from the Ontario Neurodegenerative Disease Research Initiative (ONDRI) for which overall methodology has been previously described.*^27,38,39^* In addition, the amyloid-β PET-negative cognitively unimpaired controls from the Brain-Eye Amyloid Memory study (BEAM)*^40^* underwent the same ONDRI harmonized assessments. The primary analyses compared baseline data across ten diagnostic groups. Secondary cross-sectional and longitudinal analyses were restricted to the ALS group. Screening for patient recruitment was conducted by board-certified clinicians (neurologists, geriatric psychiatrists, geriatricians) by applying prevailing diagnostic criteria for MCI, AD, PD, FTD, CVD and ALS at the time, as previously described.*^38^* Final ONDRI diagnoses were achieved by consensus review among relevant clinicians and were supported by clinical examination, clinical history, MRI and exclusionary blood work.

Key general inclusion criteria were no history of other underlying disease or substance abuse, fluency in English, Montreal Cognitive Assessment (MoCA) score ≥18 (≥14 for FTD), ≥8 years education, ability to walk and having a reliable study partner. The MCI group included patients with single- or multi-domain amnestic MCI inferred due to AD. The AD group included patients with predominantly typical AD and few atypical AD cases, the latter defined as involvement of non-memory neuropsychological domains at initial presentation, or development of other symptoms suggesting a likely co-diagnosis shortly after initial amnestic presentation. The PD-CU, PD-MCI and PD-D groups (cognitively classified according to the Movement Disorder Society Task Force Level II guidelines) included patients all initially presenting the typical motor syndrome responsive to levodopa, with no initial diagnosis of dementia with Lewy bodies. The FTD group included patients diagnosed with corticobasal syndrome, semantic dementia, progressive nonfluent aphasia, progressive supranuclear palsy and behavioral variant FTD. The CVD-CU (MoCA score ≥26) and CVD-I (MoCA score <26) groups included patients with a mild to moderate ischemic stroke event >3 months preceding recruitment, confirmed by neuroradiologists on CT/MRI; and excluded patients with history of cognitive impairment before the stroke event. The ALS group included patients with bulbar or limb onset.

Data sets are available by request from qualified researchers through *Brain-CODE*. Protocols were approved by research ethics boards at participating institutions across Ontario, Canada. Written informed consent was provided by patients before participation in accordance with the Declaration of Helsinki.

### Plasma biomarkers and genotyping

Blood samples were collected once at baseline by Lifelabs Medical Laboratory Services according to standard operating procedures and shipped unfrozen (on ice) to the ONDRI Biobank to be processed within 24 hours. *APOE* ε4 genotype, presented linearly as allele dosage (0/1/2), was determined using the ONDRISeq sequencing panel following DNA isolation from blood.*^41^* Plasma was isolated by centrifugation (15 minutes, 4°C, 2000 rpm) and stored at -80°C. Plasma biomarker quantification was then performed at the Clinical Neurochemistry Laboratory at the University of Gothenburg using ultra-sensitive Simoa immunoassays on an HD-X Analyzer (Quanterix, USA), following manufacturer specifications. Samples were run in singlicate using one batch of reagents, with two duplicate quality check samples (start/end), by laboratory technicians blinded to clinical data. Different assays were used for p-tau217 (ALZpath p-tau217 assay, ALZpath Inc., Carlsbad, CA, USA), p-tau181 (p-tau181 Advantage assay, Quanterix, Billerica, Mass, USA), Aβ42, Aβ40, GFAP and NfL (Neurology 4-plex E assay). The functional lower limit of quantification divided by two was imputed in cases of missing data due to subthreshold concentrations (<3% samples). Ratios were not computed for samples with imputed data. The biomarker data generated represents baseline levels at time of study entry. Clinical and cognitive data were obtained at baseline and in follow up annually for up to three years, in all patient groups except the ALS group who were followed every six months due to their more rapid progression. Longitudinal analyses in this study were restricted to the ALS group, as longitudinal association analyses in other diagnostic groups have been previously reported.*^27,39^*

### Statistical analyses

Plasma levels of p-tau217, p-tau181, Aβ42/40, NfL and GFAP were compared between groups using ANCOVAs adjusted for age, sex, and *APOE* ε4 allele dosage (0/1/2 alleles modelled linearly), followed by Bonferroni correction for multiple testing.

Aβ status was inferred from baseline plasma p-tau217 concentrations using published reference thresholds for the ALZpath assay.*^12^* We applied a three-range strategy, with upper and lower thresholds set to achieve 95% specificity and sensitivity for ruling in and ruling out Alzheimer’s disease, respectively, and ordinally categorised as negative, intermediate, or positive based on these thresholds. The distribution across diagnostic groups was first compared using a Kruskal-Wallis rank-sum test. Following confirmation of a significant omnibus effect, post-hoc pairwise comparisons were performed using Dunn’s test with pairwise Bonferroni correction. As a sensitivity analysis, the effect of the classification threshold on inferred Aβ positivity was examined using binomial logistic regression. Separate models were fit for each threshold and the probability of inferred Aβ positivity was estimated for each diagnostic group relative to HC, with Bonferroni correction applied across the nine group-versus-HC comparisons.

Within the ALS group, associations between baseline plasma biomarker concentrations and disease-specific clinical characteristics were examined. For measures with only a single timepoint, ordinary least-squares regressions were fit for each of the five baseline plasma biomarkers against upper motor neuron (UMN) clinical burden, lower motor neuron (LMN) clinical burden, and disease duration (months since symptom onset). UMN burden reflected involvement across three regions (bulbar, cervical, and lumbosacral), whereas LMN burden reflected involvement across four regions (bulbar, cervical, thoracic, and lumbosacral). Each cross-sectional model took the form:

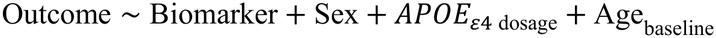

where Biomarker denotes the baseline plasma concentration and *Age_baseline_* denotes the participant’s age at the baseline visit for that outcome measure.

For measures with multiple timepoints within the ALS group, longitudinal changes in the Amyotrophic Lateral Sclerosis Functional Rating Scale-Revised (ALSFRS-R) total score, the MoCA total score, and five composite neuropsychological domains (attention and working memory, executive function, language, verbal memory, and visuospatial function) derived identically to Sanchez et al.,*^39^* were modelled with linear mixed-effects regression. MoCA was treated as a global cognitive screening measure and distinct from composite domain scores derived from individual neuropsychological tests. The ALSFRS-R model was specified as:

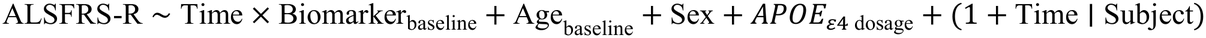

The MoCA model was specified as:

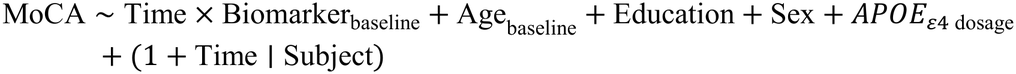

The neuropsychological domain models were specified as:

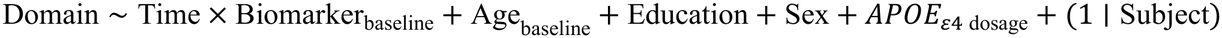

The notation × refers to both the main effect of time and the interaction terms. The domain models lack a random effect for time due to model overparametrization issues and only cleanly converged once the random effects structure was simplified to only include a random intercept per subject. Bonferroni correction was applied across the five biomarkers tested.

Receiver operating characteristic-area under the curve (ROC-AUC) analyses assessed the discriminatory capacity of plasma biomarkers and combination ratios across all diagnostic groups. Comparisons meeting prespecified criteria (AUC > 0.80) were retained for figure presentation.

All statistical analyses were performed in R version 4.4.

## Results

### Study sample characteristics

A total of 551 participants were included in this study (Table 1), of which 506 were patients diagnosed with predominantly sporadic neurodegenerative diseases (4 ALS cases had *C9orf72* hexanucleotide repeat expansions [>60]) or cerebrovascular disease and 45 were controls, previously described extensively.*^27,38,39^*

**Table 1.** Study sample characteristics across diagnostic groups.

| Diagnostic Group | HC | MCI | AD | PD-CU | PD-MCI | PD-D | FTD | CVD-CU | CVD-I | ALS | Statistic<sup>a</sup> | P value | Effect Size<sup>b</sup> |
|---|---|---|---|---|---|---|---|---|---|---|---|---|---|
| N | 45 | 85 | 41 | 48 | 52 | 26 | 53 | 86 | 75 | 40 |  |  |  |
| Age, years | 66.7 (6.0) | 70.6 (8.3) | 71.8 (8.0) | 65.9 (5.8) | 68.1 (5.8) | 69.3 (7.1) | 67.8 (7.1) | 67.8 (6.8) | 70.8 (7.8) | 62.0 (8.7) | F[9, 541] = 7.42 | **<0.001** | η² = 0.11 |
| Age range, years | 51.2–76.9 | 53.4–87.2 | 54.4–87.8 | 55.3–79 | 55.1–81.7 | 56.7–84.4 | 49.7–80.9 | 55–85.4 | 55.2–84.5 | 40.1–77.2 |  |  |  |
| Sex, % women | 75.6 | 47.1 | 41.5 | 27.1 | 21.2 | 23.1 | 35.8 | 32.6 | 30.7 | 40.0 | χ2[9] = 44.38 | **<0.001** | ω = 0.25 |
| *APOE* ε4, % carriers | 22.7 | 41.2 | 65.9 | 18.8 | 23.5 | 42.3 | 37.7 | 18.6 | 32.0 | 32.5 | χ2[9] = 41.17 | **<0.001** | ω = 0.24 |
| Education, years | 16.3 (2.0) | 15.4 (3.0) | 14.8 (3.2) | 15.8 (2.6) | 15.2 (2.9) | 15.4 (2.4) | 13.9 (2.7) | 14.6 (3.0) | 14.6 (2.8) | 13.8 (2.9) | F[9, 541] = 3.87 | **<0.001** | η² = 0.06 |
| MoCA score | 28.1 (1.4) | 23.5 (2.7) | 20.9 (2.8) | 27.5 (1.9) | 24.9 (2.5) | 24.8 (2.6) | 21.5 (3.9) | 27.5 (1.3) | 22.6 (2.1) | 25.5 (2.8) | F[9, 540] = 59.36 | **<0.001** | η² = 0.50 |

Continuous variables: mean (SD). Sex and *APOE* ε4: percentage of group. AD = Alzheimer's disease; ALS = amyotrophic lateral sclerosis; *APOE* = apolipoprotein E; CVD-CU = cerebrovascular disease, cognitively unimpaired; CVD-I = cerebrovascular disease, cognitively impaired; FTD = frontotemporal dementia; HC = cognitively unimpaired controls; MCI = mild cognitive impairment; MoCA = Montreal Cognitive Assessment; PD-CU = Parkinson's disease, cognitively unimpaired; PD-D = Parkinson's disease with dementia; PD-MCI = Parkinson's disease with mild cognitive impairment; SD = standard deviation.
<sup>a</sup> F statistic from one-way analysis of variance for continuous variables; χ² statistic for categorical variables. P values correspond to these omnibus tests.
<sup>b</sup> η² = eta-squared for continuous variables; ω = bias-corrected Cramér's V for categorical variables.

Significant differences across diagnostic groups were observed for all clinical and demographic characteristics (Table 1). Age varied significantly across the study sample (F[9, 541] = 7.42, P < 0.001, *η*^2^ = 0.11), with the ALS group being the youngest on average (mean [SD] = 62.0 [8.7] years) and the AD group being the oldest (mean [SD] = 71.8 [8.0] years). The proportion of female participants differed significantly by group (*χ*^2^[9] = 44.38, P < 0.001, *ω* = 0.25), with the control group showing the highest percentage (75.6%) and the PD-MCI group showing the lowest (21.2%). As expected, the prevalence of *APOE ε*4 carriers differed significantly across groups (*χ*^2^[9] = 41.17, P < 0.001, *ω* = 0.24), peaking in the AD group (65.9%) followed by PD-D (42.3%), and being lowest in the CVD-CU group (18.6%). Years of education also differed significantly (F[9, 541] = 3.87, P < 0.001, *η*^2^ = 0.06), with controls having the highest average level (16.3 [2.0] years) and the ALS group having the lowest (13.8 [2.9] years). Lastly, cognitive performance measured by the MoCA showed highly significant differences (F[9, 540] = 59.36, P < 0.001, *η*^2^ = 0.50), with the AD group presenting the lowest mean score (20.9 [2.8]) and controls presenting the highest (28.1 [1.4]).

### Biomarker levels

Plasma levels of p-tau217, p-tau181, Aβ42/40, NfL and GFAP were compared between groups with ANCOVAs adjusted for age, sex, and *APOE* ε4 allele dosage modeled in a continuous fashion, followed by pairwise post-hoc comparisons Bonferroni-corrected for multiple testing (Figure 1). Group effects were found for p-tau217 (F[9, 526] = 16.47, P < 0.001), p-tau181 (F[9, 528] = 4.01, P < 0.001), GFAP (F[9, 535] = 6.64, P < 0.001) and NfL (F[9, 535] = 24.22, P < 0.001) but not for Aβ42/40 (F[9, 508] = 1.57, P = 0.12). Compared to HC, levels of p-tau217 were elevated only in patients with AD (t[526] = -6.83, P < 0.001) or ALS (t[526] = -4.74, P < 0.001); levels of p-tau181 and GFAP were elevated only in patients with AD (t[528] = -5.01, P < 0.001; t[535] = -5.20, P < 0.001); levels of NfL were elevated only in patients with ALS (t[535] = -12.46, P < 0.001) or FTD (t[535] = -3.67, P = 0.012). Full ANCOVA and *post-hoc* results are provided in Supplementary Table S1 and S2, respectively.

**Figure 1:**
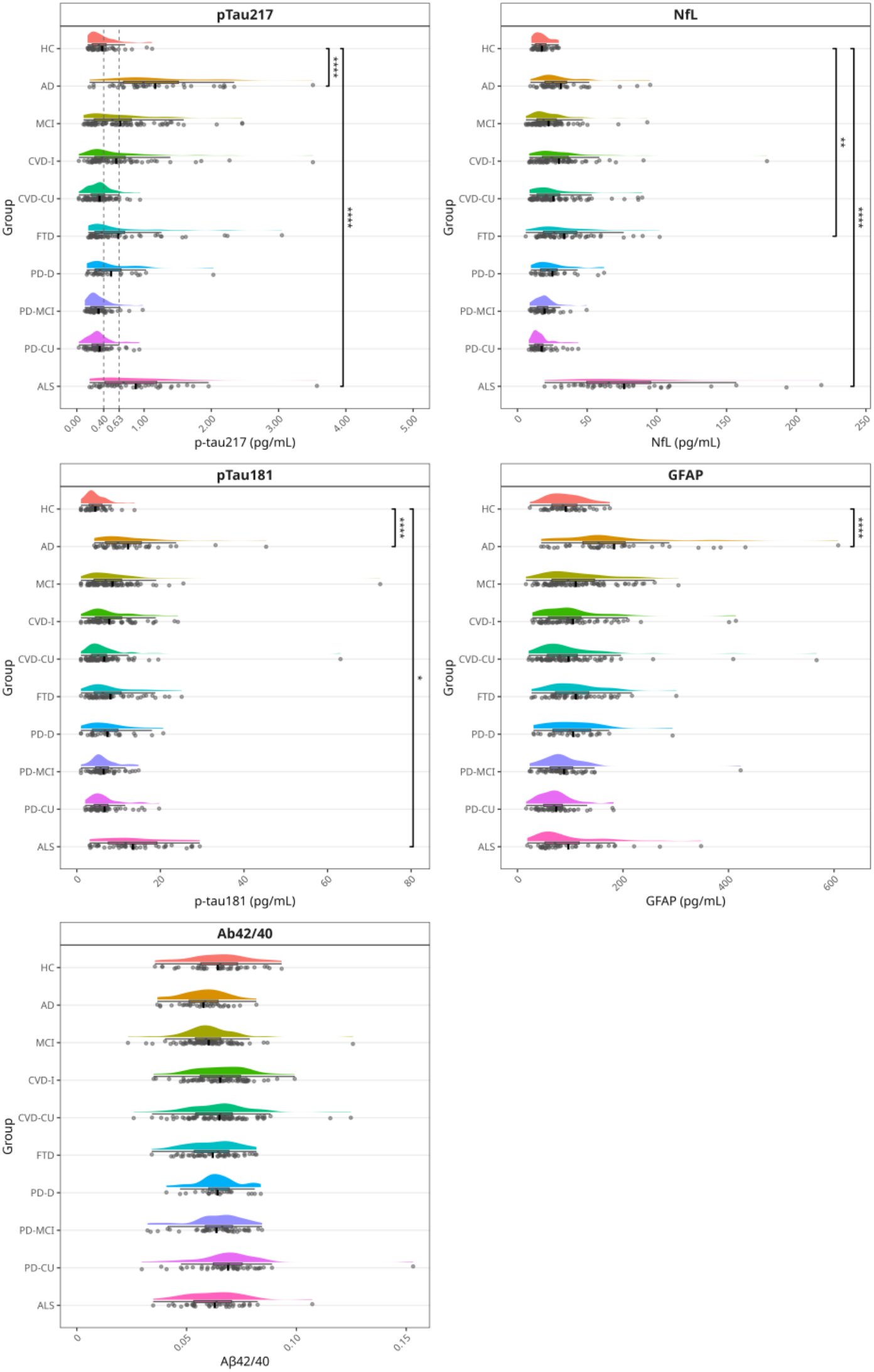
Comparative levels of plasma biomarkers. Raincloud plot line and error bars represent mean and 95 percent confidence interval. Bonferroni-corrected comparisons to controls (HC) are shown. Some data points were truncated for display purposes: in the AD group, one p-tau217 data point (7.31 pg/mL), two p-tau181 data points (113 pg/mL, 284.4 pg/mL) and one *Aβ*42/40 data point (0.204 pg/mL); in the MCI group, one GFAP data point (1248 pg/mL) and one NfL data point (309 pg/mL). For the p-tau217 plot, vertical dashed lines represent the lower and upper thresholds for inferred amyloid-β positivity as per Ashton et al.*^12^* * P < 0.05, ** P < 0.01, *** P < 0.001, **** P < 0.0001.

### Inferred amyloid-β positivity

Inferred Aβ status was classified according to published plasma p-tau217 reference thresholds*^12^* and is depicted in Figure 2 A (frequency counts and percentages are provided in Supplementary Table S3). The proportion classified as Aβ positive increased stepwise from HC (9.3%) to MCI (40%) to AD (82.5%). Other notable observations relating to inferred Aβ positive proportions included ALS (55%), PD-D (30.8%), and FTD (28.3%). PD-CU (8.3%), PD-MCI (5.9%), and CVD-CU (3.6%) exhibited the lowest proportions of positivity.

**Figure 2:**
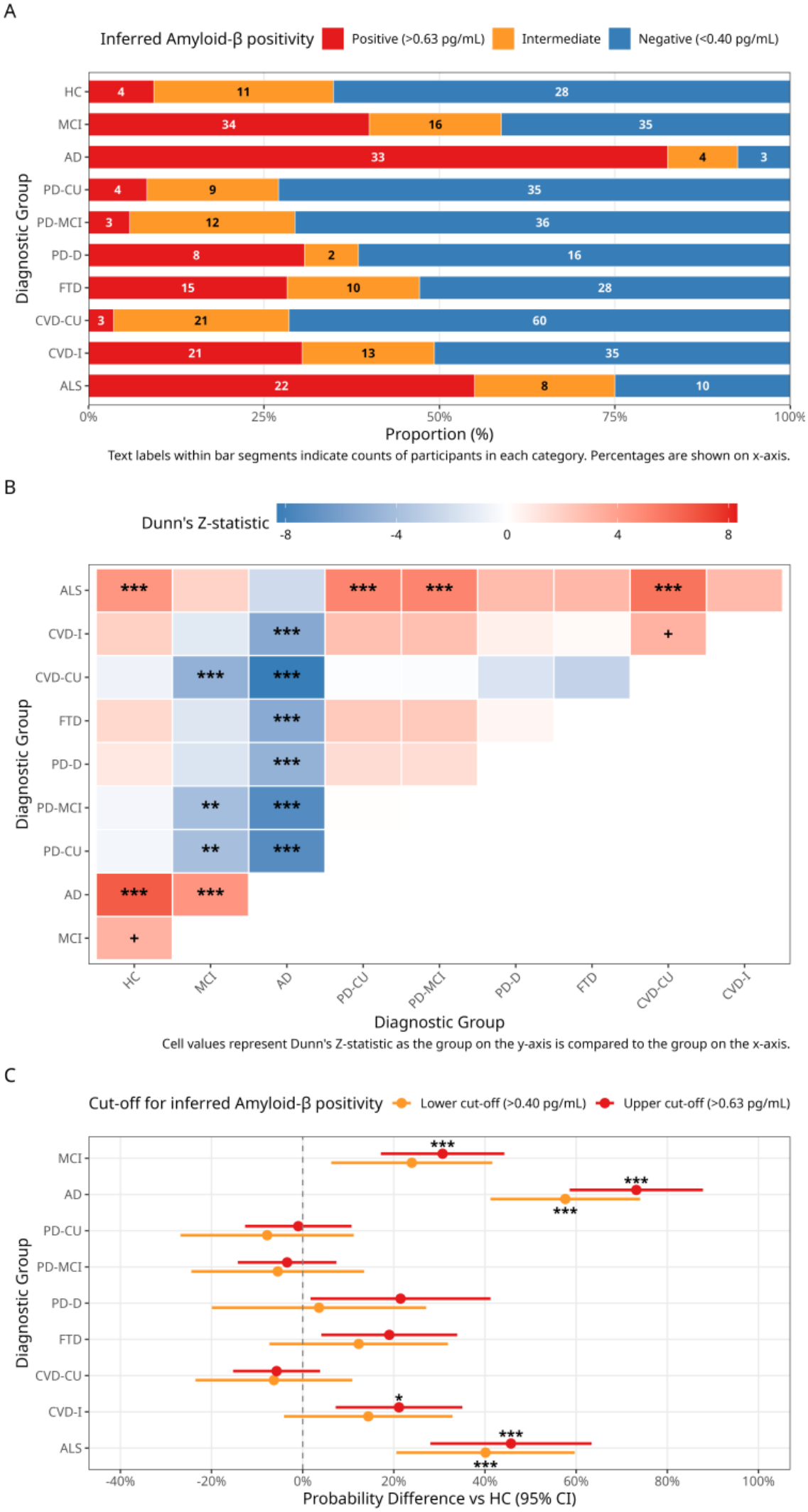
Inferred Aβ positivity classification by diagnostic group. (**A**) Stacked proportions of Aβ negativity, intermediate, and positivity by diagnostic group using published p-tau217 reference thresholds.*^12^* Numbers within coloured bar segments indicate participant counts; percentages are shown on the x-axis. (**B**) Upper triangle heatmap of all pairwise Dunn’s test comparisons (Z-statistic). Overall Kruskal-Wallis test: *χ*^2^(9) = 116.22, P < 0.001. Significance after Bonferroni correction: + P < 0.06, * P < 0.05, ** P < 0.01, *** P < 0.001. (**C**) Sensitivity analysis of Aβ positivity probability relative to HC using lower (p-tau217 > 0.40 pg/mL) and strict (p-tau217 > 0.63 pg/mL) thresholds, estimated by logistic regression with Bonferroni correction.

A Kruskal-Wallis test was used to assess overall group differences, which was significant (*χ*^2^[9] = 116.22, P < 0.001). *Post-hoc* Dunn’s tests (Figure 2 B) confirmed that, relative to HC, mean rank scores of Aβ positivity were significantly elevated in MCI (Z = 3.24, P = 0.011), AD (Z = 6.62, P < 0.001), and ALS (Z = 4.40, P < 0.001). Prior to multiple comparison correction, CVD-I showed an uncorrected trend (Z = 2.00, P = 0.045 uncorrected). Complete comparisons are provided in Supplementary Table S4.

As a sensitivity analysis, the effect of the classification threshold on the probability of inferred Aβ positivity relative to HC was examined using logistic regression under both a lower (p-tau217 > 0.40 pg/mL) and upper (p-tau217 > 0.63 pg/mL) threshold (Figure 2 C). Under the upper threshold, elevated probability of a positive classification relative to HC was observed for MCI (+31%, 95% CI 12-50%, Z = 4.44, P < 0.001), AD (+73%, 95% CI 52-94%, Z = 9.81, P < 0.001), ALS (+46%, 95% CI 21-71%, Z = 5.06, P < 0.001), and CVD-I (+21%, 95% CI 1-41%, Z = 2.98, P = 0.026). Both MCI and CVD-I lose significance under the lower threshold. PD-D and FTD did not differ significantly from HC under either threshold (upper: PD-D, P = 0.30; FTD, P = 0.11). Full logistic regression probability difference results are provided in Supplementary Table S5.

### ALS biomarker and clinical associations

Clinical characteristics specific to the ALS group are displayed in Table 2. Associations between baseline plasma levels of p-tau217, p-tau181, Aβ42/40, NfL, and GFAP and baseline clinical features (LMN burden, UMN burden, and disease duration) were evaluated using linear regression models adjusted for age, sex, and *APOE ε*4 allele dose. None of these baseline associations remained statistically significant after Bonferroni correction. A potential trend was observed between higher baseline plasma NfL levels and greater LMN burden scores (*β* = 0.0087, partial *R*^2^ = 0.14, P = 0.024 uncorrected, P = 0.11 Bonferroni corrected).

**Table 2.** Clinical and neuropsychological characteristics of the ALS sample.

| Clinical Characteristic | Value |
|---|---|
| N | 40 |
| Site of onset |  |
| Bulbar | 5 (12.5) |
| Spinal | 35 (87.5) |
| Disease duration, months | 25.7 (14.5) |
| ALSFRS, score (max 48) | 38.5 (4.6) |
| ALSFRS decline rate per month, score | -0.5 (0.3) |
| LMN burden (max 4 regions) | 3.1 (0.8) |
| UMN burden (max 3 regions) | 2.0 (0.8) |
| Neurocognitive assessments |  |
| MoCA, total score (max 30) | 25.5 (2.8) |
| Attention & working memory, z score | 0.09 (0.64) |
| Executive function, z score | 0.16 (0.56) |
| Language, z score | 0.09 (0.51) |
| Verbal memory, z score | 0.06 (0.63) |
| Visuospatial, z score | 0.24 (0.43) |

Categorical variable (site of onset) is expressed as n (%). Continuous clinical characteristics, MoCA total score, and neuropsychological domain z-scores are expressed as mean (SD). ALS = amyotrophic lateral sclerosis; ALSFRS = Amyotrophic Lateral Sclerosis Functional Rating Scale; LMN = lower motor neuron; MoCA = Montreal Cognitive Assessment; SD = standard deviation; UMN = upper motor neuron.

The longitudinal relationship between baseline plasma biomarkers and clinical progression was evaluated against changes in ALSFRS-R, MoCA scores, and neuropsychological domain z-scores over time. No longitudinal associations survived strict multiple comparison correction. A significant interaction prior to correction was observed between baseline NfL and time such that elevated levels of NfL were associated with a faster rate of ALSFRS-R clinical decline (*β* = - 0.099, P = 0.012 uncorrected, P = 0.059 Bonferroni corrected). Complete association results are provided in Supplementary Tables S6-S9.

One of the ALS cases came to autopsy, which confirmed a final neuropathologic diagnosis of Amyotrophic Lateral Sclerosis with predominantly lower motor neuron degeneration and Alzheimer’s type neuropathologic change (ADNC), A1B1C1. See Supplementary Material section *ALS Neuropathology Case* for clinical and pathological details of the case.

### ROC-AUC analysis

Under a discriminatory threshold of AUC > 0.80, the pairwise ROC-AUC analysis across all clinical category combinations highlighted performant single biomarkers and combination ratios (Figure 3).

**Figure 3:**
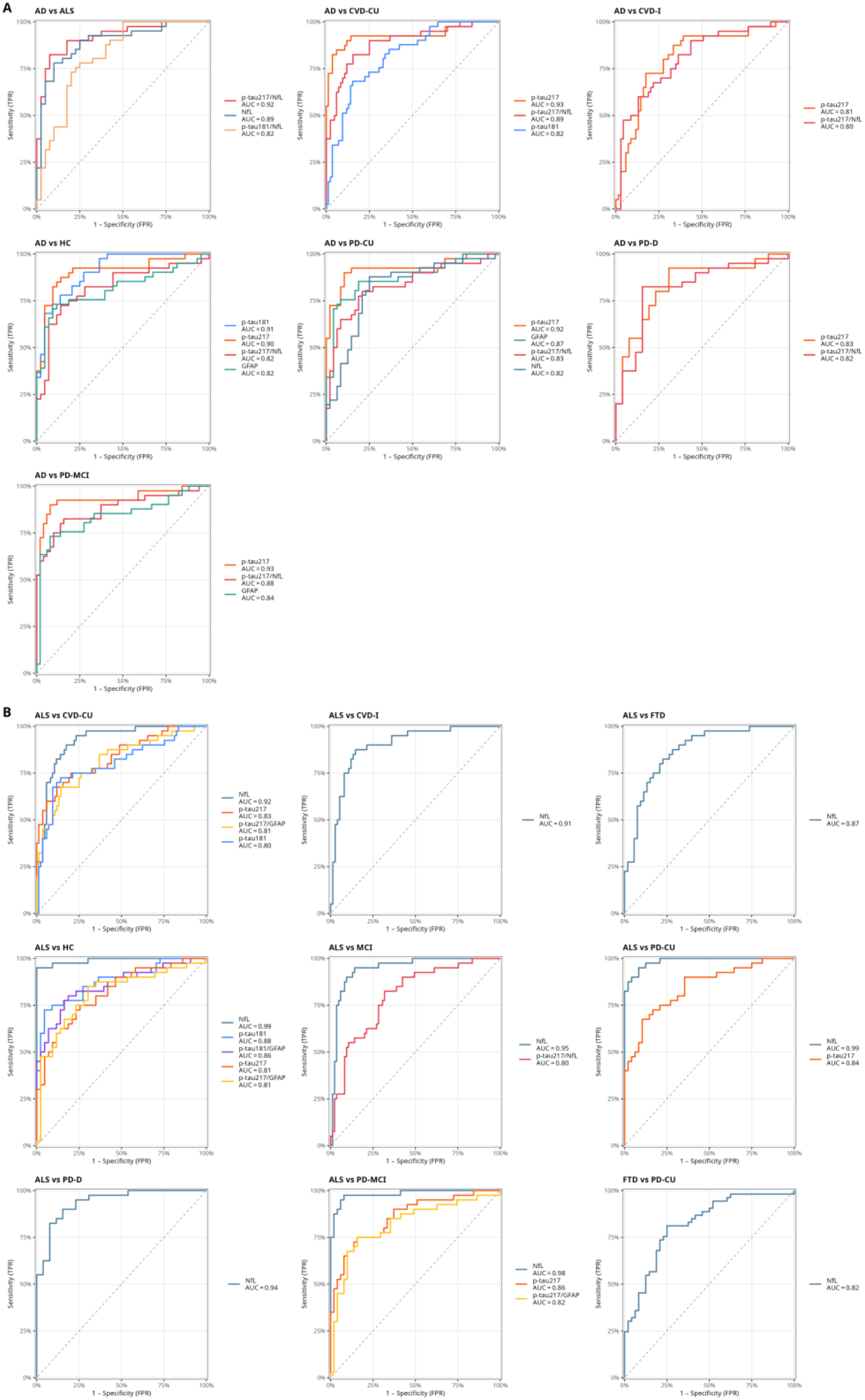
Receiver operating characteristic (ROC) curves for plasma biomarkers and combination ratios in distinguishing diagnostic categories thresholded at an area under the curve (AUC) > 0.80 for at least one biomarker per comparison. (**A**) Comparisons separating the AD group from other diagnostic groups. (**B**) Comparisons separating the ALS group from other groups, along with a comparison separating FTD from PD-CU.

Several biomarkers or combinatoric ratios surpassed the AUC threshold discriminating AD from other diagnoses (Figure 3, Panel A), most prominently by p-tau217 when differentiating from PD groups (PD-CU: AUC = 0.92, PD-MCI: AUC = 0.93, PD-D: AUC = 0.83) and cerebrovascular disease groups (CVD-CU: AUC = 0.93, CVD-I: AUC = 0.81). Both p-tau181 (AUC = 0.91) and p-tau217 (AUC = 0.90) separated AD from HC and performed better than their ratios with NfL or compared to GFAP (AUC = 0.82). Notably, the combination ratio p-tau217/NfL showed even better performance of differentiating ALS from AD (AUC = 0.92) compared to NfL alone (AUC = 0.89).

NfL emerged as a consistent biomarker for identifying ALS (Figure 3, Panel B), with high diagnostic accuracy against controls (AUC = 0.99), Parkinson’s disease groups (PD-CU: AUC = 0.99; PD-MCI: AUC = 0.98; PD-D: AUC = 0.94), Alzheimer’s disease impairment groups (MCI: AUC = 0.95; AD: AUC = 0.89), cerebrovascular disease groups (CVD-CU: AUC = 0.92; CVD-I: AUC = 0.91), and FTD (AUC = 0.87). It was also observed that NfL provided some discriminatory power between FTD and PD-CU cases (AUC = 0.82). A complete overview of the results is provided in Supplementary Table S10.

## Discussion

In this study we evaluated plasma p-tau217, p-tau181, Aβ42/40, NfL, and GFAP in a deeply phenotyped study sample across ten clinically defined diagnostic groups. Compared to controls, p-tau217 was significantly elevated in AD and ALS, whereas p-tau181 was elevated only in AD. Inferred Aβ positivity relative to HC increased as expected in MCI and AD, as well as in ALS and CVD-I, the lattermost of which points to potential AD or cerebral amyloid angiopathy (CAA) co-pathology. While not meeting the threshold for statistical significance, approximately 30% of PD-D and 28% of FTD cases also had inferred Aβ positivity indicating potential co-pathology in PD-D and possibly FTD, or misdiagnosis in the case of FTD. This is consistent with post-mortem evidence that additional disease- or age-associated pathologies frequently accompany the expected primary pathology across neurodegenerative diseases.*^13^* The p-tau217 elevations within the ALS group also prompted investigation into potential correlation with clinical and cognitive features, although no statistically significant associations were found. This is in contrast to cross-disease cognitive and neuroimaging associations identified between p-tau species in AD, PD, and CVD groups in this sample where baseline p-tau species correlated with both baseline cognitive performance and longitudinal cognitive decline,*^39^* as well as progressive cerebral atrophy and white matter hyperintensity burden.*^27^* The increase in p-tau217 observed in both AD and ALS groups impaired differentiation between these two diseases using this biomarker alone. However, pairing p-tau217 with NfL enabled superior discrimination between ALS and AD compared with any single biomarker.

As illustrated elsewhere,*^17^* recognizing the potentially high prevalence of clinically relevant AD existing as a co-pathology is paramount across a range of presentations, and thus reliance on AD biomarkers used in isolation may not be sufficient to achieve adequate primary aetiological categorisation in these instances. Of course, diagnostic uncertainty rarely exists in standard clinical practice between AD and ALS, although the contribution of AD co-pathology to cognitive deficits in ALS is increasingly recognised.*^42^* However, utilising plasma p-tau species in isolation for the identification of preclinical disease or even in cases lacking well-defined clinical syndromes may carry a substantial risk of diagnostic misclassification if used indiscriminately.

Plasma p-tau species have been demonstrated to be highly accurate in identifying biologically defined AD when established biomarkers are used as reference standards.*^43^* In the BioFINDER-2 cohort, p-tau181 enabled differentiation of AD from other neurodegenerative disorders, although this was a pre-selected sample in which non-AD cases were confirmed Aβ-negative via PET and ALS cases were not represented.*^11^* In line with these studies, we also found elevated levels of p-tau217 and p-tau181 in AD relative to cognitively unimpaired controls (Figure 1; Supplementary Table S1). However, elevations in blood phosphorylated tau species, including p-tau217, have also been reported in ALS by several independent groups.*^28–30,44^* The elevation of plasma p-tau217 in ALS appears to be somewhat counterintuitive given that this disease is primarily characterised by TDP-43 pathology, and CSF phosphorylated tau is also consistently non-elevated,*^31,45^* suggesting that this signal might not necessarily originate from the CNS. Elevated plasma p-tau181 was initially shown to correlate with lower motor neuron dysfunction and greater spinal cord neuronal loss in clinicopathologically defined ALS cases, but not with motor cortex pathology.*^28^* Furthermore, they observed no correlations between plasma p-tau181 and AD-specific CSF biomarkers (i.e., p-tau181, Aβ42), as well as CSF biomarkers of neurodegeneration (i.e., total-tau, NfL).*^28^* These findings were further substantiated by another group additionally characterised with neurophysiology, and a small number of Spinal Muscular Atrophy (SMA) patients studied also exhibited similar elevations.*^29^* Both studies similarly observed no correlation with cognitive impairment in ALS and those with combined FTD/ALS in fact exhibited lower levels of p-tau181.*^28,29^* The conclusions were that the elevation in plasma p-tau species in ALS may have largely originated extra-cerebrally. More recently, p-tau217 and p-tau181 have been directly found in atrophic muscle fibres of ALS cases.*^44^* Interestingly this was also observed in the control sample, and correlated with age, additionally indicating a possible senescence-related effect, such as sarcopenia.

Our findings align with this interpretation. Within the ALS group, no baseline biomarker showed significant associations with ALS clinical features or predicted longitudinal change in ALSFRS-R, MoCA scores, or neuropsychological domain z-scores after correction for multiple comparisons (Supplementary Tables S6-S9). The single autopsy-confirmed ALS case in our study sample (Supplementary Material section *ALS Neuropathology Case*) exhibited only low-level ADNC (A1B1C1) which was insufficient to account for the observed p-tau217 elevation. Antemortem validation studies have shown that significant p-tau217 elevations measured with the ALZpath assay do not occur in participants with none/low ADNC.*^46^* While marginal changes of p-tau217 can be exhibited at early stages of Aβ accumulation, this is typically observed only once centiloid values exceed approximately 35 on Aβ-PET,*^8,47^* further arguing against incidental preclinical AD as a sufficient explanation for the magnitude of elevation observed in our ALS group, at least in this case.

Alterations in tau metabolism in ALS have been observed,*^48^* given sparse evidence of increased tau phosphorylation in both the upper and lower motor neurons of sporadic ALS cases in the absence of clinically apparent cognitive or behavioural impairment.*^49^* It is not clear if these changes play a direct pathogenic role or merely reflect a secondary neuronal stress response, and in fact tau phospho-epitopes differing to those examined in AD have been implicated in this context (pThr175, pSer214, pSer396, pSer404).*^48^*Moreover, when pathological brain tau deposition was systematically evaluated in a moderate sized ALS/FTD neuropathological cohort, excluding Frontotemporal Lobar Degeneration-Tau (FTLD-Tau) cases, this was found only to be limited in extent and in most instances considered more likely to represent co-incident AD or Primary Age-Related Tauopathy (PART).*^50^* Crucially, PART has not been associated with elevations in plasma p-tau species.*^19^*

The predominating tau isoform in the peripheral nervous system (so called “big tau”) is nearly double the molecular mass of those lower molecular weight (LMW) tau isoforms expressed in the brain, which is thought to be critical in regulating axonal transport in large projecting axons.*^51^* This is due to additional inclusion of exon 4a of *MAPT* which generates a high-molecular weight isoform.*^52^* Crucially, currently available assays differ in their ability to distinguish between the lower and higher-molecular weight isoforms, depending on the detection antibody employed which may or may not target epitopes common to both.*^53^* A group has now demonstrated that LMW-specific assays show less marked elevations in ALS and could discriminate AD from ALS with higher accuracy as opposed to non-LMW-specific assays, including the ALZpath and Quanterix Simoa assays we have utilised here.*^54^* Therefore, this seems to highlight the potential for peripherally derived p-tau species to confound detection of the brain-derived plasma fraction. This of course poses significant questions about these phosphorylation epitopes serving as biomarkers of CNS pathology, such as AD-specific processes. It is not presently clear if this observation is restricted to ALS or may be a consideration in other neuromuscular disorders as well. Notably, elevations in blood p-tau181 and p-tau217 have now likewise been reported in patients with systemic amyloidosis and associated polyneuropathy.*^55^*

In our study, inferred Aβ positivity based on published p-tau217 thresholds increased stepwise from controls through MCI to AD (Figure 2 A), confirming the thresholds set out by Ashton et al.*^12^* were applicable to our sample. Of note, inferred Aβ positivity rates in the AD group were elevated compared to most other diagnostic groups other than ALS (Figure 2 B). In our sensitivity analysis assessing the impact of the lower versus upper cut-off thresholds for inferred Aβ positivity, the latter criteria yielded an increase in the rate of inferred Aβ positivity in the CVD-I group (Figure 2 C; Supplementary Table S5). This finding could be consistent with the well-established co-occurrence of Alzheimer’s and cerebrovascular disease in later-life cognitive impairment, as demonstrated by community-based autopsy series*^56^* and PET imaging.*^15^* It is also increasingly recognised that their effects on cognition may not be merely summative but may be mutually reinforcing.*^57,58^* Thus, assigning the correct correspondence of prospective biomarkers to each of these disease mechanisms independently, when they are not likely to be fully dichotomous, poses a particular challenge. Both p-tau181 and p-tau217 have been found to correlate with white matter hyperintensity volume*^59–61^* and radiological indices of CAA,*^60,62,63^* and enabled adequate distinction between deep perforator arteriopathy, CAA and AD in one cohort.*^20^* Specifically, AD patients had highest levels of p-tau217, followed by CAA and then by those with deep perforator arteriopathy.*^20^* However, most of these studies lacked definitive neuropathological examination and therefore it is unclear to what extent these associations are exclusively mediated by CAA, or how to isolate the contributions of parenchymal and vascular β-amyloidosis to an observed biomarker profile in this context given the intersecting pathobiologies of CAA, AD and cerebrovascular disease.*^16^* While there strictly was no statistically significant difference in group-wise comparisons between PD-D and FTD in this study sample, there was still a notable proportion in these groups (over 25%) who reached the upper threshold for inferred Aβ positivity considering our sample size. This once more serves to illustrate the role of concomitant AD neuropathology even in prototypical clinical presentations.*^14,17^* The high incidence of reciprocal co-pathology between AD and Lewy body disease, including PD-D, is increasingly recognized,*^14^* and other investigators examining blood biomarkers in this setting have shown that p-tau species can help delineate this.*^25,26^*

Our ROC-AUC analyses reinforce the enhanced diagnostic performance of a combined biomarker strategy, with the ratio of p-tau217 and NfL providing the highest discriminatory accuracy between ALS and AD compared to any single biomarker alone (Figure 3 A; Supplementary Table S10) and the utility of this ratio in differentiating AD from frontotemporal lobar degeneration has already been established.*^64^* Notwithstanding this, our results did recapitulate diagnostic performance of standalone biomarker use across other comparisons. p-tau217 alone demonstrated strong performance (AUC > 0.8) in differentiating AD from cognitively unimpaired controls, Parkinson’s disease variants, and cerebrovascular disease groups (Figure 3 A). Similarly, NfL alone was the top performing biomarker in differentiating ALS from cognitively unimpaired controls, Alzheimer’s & MCI variants, Parkinson’s disease variants, FTD, and cerebrovascular disease groups (Figure 3 B). While NfL elevation is an ubiquitous finding across neurodegenerative diseases as a marker of neuroaxonal injury,*^65^* precluding its use as a specific etiological marker, it has been shown to exhibit high diagnostic performance in distinguishing ALS from mimic presentations, demonstrating superiority over p-Tau181 and GFAP in this context.*^66^*

Strengths of this study include the harmonised recruitment of participants with different neurodegenerative diseases across sites, the breadth of diagnostic groups, and the use of a single assay platform. Several limitations apply. Definitive confirmation of Aβ status was not obtained via CSF or PET imaging, and inferred positivity therefore relied on published plasma p-tau217 thresholds. The study sample largely lacked neuropathological data, and analytes were examined in one biofluid only, necessitating indirect inference regarding their tissue source; future studies employing LMW-specific assays for p-tau quantification will be informative. Clinical phenotyping without biomarker-confirmed molecular subtyping means that occult AD co-pathology in non-AD groups cannot be excluded.

In summary, this comprehensive analysis of conventionally available plasma biomarkers across the full spectrum of neurodegenerative and cerebrovascular disease highlights several important diagnostic implications regarding their real-world implementation. Our results show that p-tau181 and p-tau217 species do not appear to be uniquely specific to AD, emphasising the importance of correctly understanding the precise dynamics of pathological tau phosphorylation across differing neurodegenerative disorders.*^67^* This underscores the need for identification of more definitive biomarkers for each neurodegenerative disease, both in terms of a truly CNS originating p-tau isoform*^68^* and one that specifically corresponds to neurofibrillary tangles of AD,*^69,70^* if the utility of Tau PET imaging is to be fully emulated.*^71^* Additionally, we still do not possess adequate biomarkers for other common neurodegenerative disorders when co-pathology is often found to be the norm.*^13^*

If molecular diagnostic testing in plasma is proposed to supplant traditional neuropathological confirmation, molecular neuroimaging and CSF examination, or even become the prevailing method of diagnosis, much of the complementary information provided by direct tissue examination is elided. Accordingly, to achieve comparable diagnostic capability, it is evident that a broader panel of biomarkers will be necessitated. This is particularly pertinent as only one neuropathological process (i.e., AD neuropathologic change) can be reliably detected *in vivo* by currently available plasma biomarker methods and is one that exists on a neurodegenerative disease continuum. Thus, the presence of all other possibly contributory proteinopathies can only be presently inferred by clinical criteria alone antemortem, although promising seed amplification assays for alpha-synuclein and 4R/4R tauopathies are emerging.*^72,73^* This has obvious ramifications for the implementation of selective therapeutics. Therefore, it is likely that panel examination of all potentially relevant molecular entities will be required before these biomarkers can be meaningfully incorporated into modern clinical practice for truly comprehensive patient profiling. Nevertheless, our study findings show that when currently available plasma constituents are serviced in a judicious manner in well characterized populations (i.e. recognising the caveats of their use),*^10^* they can serve as helpful diagnostic adjuncts in a clinical setting.

## Supporting information

Supplementary material accompanies this preprint as a separate file

## Data Availability

Data sets are available by request from qualified researchers through Brain-CODE.

## Data Availability

Curated data from the ONDRI cohort are available through controlled release on Brain-CODE (www.braincode.ca).

## Supplementary Material

Supplementary material accompanies this preprint as a separate file.

