## Supplementary material accompanies this preprint as a separate file for "Plasma phosphorylated tau species across neurodegenerative and cerebrovascular diseases: An ONDRI study"

### Supplementary Materials

#### Supplementary Tables

##### Plasma biomarker levels

***Table S1.*** *Type III ANCOVA model summaries for each plasma biomarker. All models adjusted for age, sex, and APOE ε4 allele dosage.*

| Term | df | SSR | MSS | F | P-value |
|---|---|---|---|---|---|
| p-tau217 | | | | | |
| Group | 9 | 38.692 | 4.299 | 16.47 | **<0.001** |
| Age | 1 | 2.648 | 2.648 | 10.143 | **0.002** |
| Sex | 1 | 0.138 | 0.138 | 0.53 | 0.467 |
| *APOE* ε4 allele dose | 1 | 3.021 | 3.021 | 11.572 | **<0.001** |
| Residuals | 526 | 137.296 | 0.261 | | |
| p-tau181 | | | | | |
| Group | 9 | 7053.397 | 783.711 | 4.011 | **<0.001** |
| Age | 1 | 117.97 | 117.97 | 0.604 | 0.438 |
| Sex | 1 | 251.673 | 251.673 | 1.288 | 0.257 |
| *APOE* ε4 allele dose | 1 | 172.777 | 172.777 | 0.884 | 0.347 |
| Residuals | 528 | 103174.224 | 195.406 | | |
| Aβ42/40 | | | | | |
| Group | 9 | 0.003 | 3.36e-4 | 1.575 | 0.120 |
| Age | 1 | 0.002 | 0.002 | 8.939 | **0.003** |
| Sex | 1 | 9.04e-4 | 9.04e-4 | 4.241 | **0.040** |
| *APOE* ε4 allele dose | 1 | 8.93e-4 | 8.93e-4 | 4.191 | **0.041** |
| Residuals | 508 | 0.108 | 2.13e-4 | | |
| NfL | | | | | |
| Group | 9 | 110599.235 | 12288.804 | 24.22 | **<0.001** |
| Age | 1 | 6842.699 | 6842.699 | 13.486 | **<0.001** |
| Sex | 1 | 1739.584 | 1739.584 | 3.429 | 0.065 |
| *APOE* ε4 allele dose | 1 | 439.621 | 439.621 | 0.866 | 0.352 |
| Residuals | 535 | 271447.687 | 507.379 | | |
| GFAP | | | | | |
| Group | 9 | 358138.529 | 39793.17 | 6.636 | **<0.001** |
| Age | 1 | 237112.575 | 237112.575 | 39.541 | **<0.001** |
| Sex | 1 | 164854.177 | 164854.177 | 27.491 | **<0.001** |
| *APOE* ε4 allele dose | 1 | 3897.035 | 3897.035 | 0.65 | 0.421 |
| Residuals | 535 | 3208171.807 | 5996.583 | | |

Type III sum of squares. *APOE* = apolipoprotein E; Aβ42/40 = amyloid-β42 to amyloid-β40 ratio; df = degrees of freedom; MSS = mean square; SSR = sum of squares; GFAP = glial fibrillary acidic protein; NfL = neurofilament light chain; p-tau181 = tau phosphorylated at threonine 181; p-tau217 = tau phosphorylated at threonine 217.

***Table S2.*** *ANCOVA post-hoc pairwise comparisons for all plasma biomarkers adjusting for age, sex, and APOE ε4 allele dosage.*

| Contrast | Estimate | SE | df | t-value | 95% CI low | 95% CI high | P-value (raw) | P-value adj. (vs HC)[a] | P-value adj. (vs all pairs)[b] |
|---|---|---|---|---|---|---|---|---|---|
| **p-tau217** | | | | | | | | | |
| HC - AD | -0.804 | 0.118 | 526 | -6.827 | -1.035 | -0.573 | **<0.001** | **<0.001** | **<0.001** |
| HC - MCI | -0.199 | 0.098 | 526 | -2.04 | -0.391 | -0.007 | **0.042** | 0.377 | 1 |
| HC - (CVD-I) | -0.176 | 0.102 | 526 | -1.73 | -0.376 | 0.024 | 0.084 | 0.757 | 1 |
| HC - (CVD-CU) | 0.032 | 0.098 | 526 | 0.331 | -0.16 | 0.224 | 0.740 | 1 | 1 |
| HC - FTD | -0.216 | 0.107 | 526 | -2.028 | -0.426 | -0.007 | **0.043** | 0.388 | 1 |
| HC - (PD-D) | -0.095 | 0.13 | 526 | -0.736 | -0.35 | 0.159 | 0.462 | 1 | 1 |
| HC - (PD-MCI) | 0.053 | 0.109 | 526 | 0.487 | -0.161 | 0.267 | 0.626 | 1 | 1 |
| HC - (PD-CU) | 0.011 | 0.11 | 526 | 0.103 | -0.204 | 0.227 | 0.918 | 1 | 1 |
| HC - ALS | -0.543 | 0.115 | 526 | -4.735 | -0.769 | -0.318 | **<0.001** | **<0.001** | **<0.001** |
| AD - MCI | 0.605 | 0.1 | 526 | 6.069 | 0.409 | 0.8 | **<0.001** | | **<0.001** |
| AD - (CVD-I) | 0.628 | 0.105 | 526 | 5.991 | 0.422 | 0.834 | **<0.001** | | **<0.001** |
| AD - (CVD-CU) | 0.836 | 0.104 | 526 | 8.08 | 0.633 | 1.04 | **<0.001** | | **<0.001** |
| AD - FTD | 0.587 | 0.11 | 526 | 5.35 | 0.372 | 0.803 | **<0.001** | | **<0.001** |
| AD - (PD-D) | 0.709 | 0.131 | 526 | 5.413 | 0.451 | 0.966 | **<0.001** | | **<0.001** |
| AD - (PD-MCI) | 0.857 | 0.112 | 526 | 7.62 | 0.636 | 1.078 | **<0.001** | | **<0.001** |
| AD - (PD-CU) | 0.815 | 0.115 | 526 | 7.086 | 0.589 | 1.041 | **<0.001** | | **<0.001** |
| AD - ALS | 0.261 | 0.121 | 526 | 2.161 | 0.024 | 0.498 | **0.031** | | 1 |
| MCI - (CVD-I) | 0.023 | 0.083 | 526 | 0.276 | -0.141 | 0.187 | 0.783 | | 1 |
| MCI - (CVD-CU) | 0.232 | 0.08 | 526 | 2.884 | 0.074 | 0.389 | **0.004** | | 0.184 |
| MCI - FTD | -0.017 | 0.09 | 526 | -0.191 | -0.194 | 0.16 | 0.849 | | 1 |
| MCI - (PD-D) | 0.104 | 0.115 | 526 | 0.902 | -0.122 | 0.33 | 0.368 | | 1 |
| MCI - (PD-MCI) | 0.252 | 0.092 | 526 | 2.735 | 0.071 | 0.433 | **0.006** | | 0.290 |
| MCI - (PD-CU) | 0.211 | 0.095 | 526 | 2.223 | 0.025 | 0.397 | **0.027** | | 1 |
| MCI - ALS | -0.344 | 0.102 | 526 | -3.377 | -0.544 | -0.144 | **<0.001** | | **0.035** |
| (CVD-I) - (CVD-CU) | 0.209 | 0.083 | 526 | 2.499 | 0.045 | 0.373 | **0.013** | | 0.574 |
| (CVD-I) - FTD | -0.04 | 0.094 | 526 | -0.429 | -0.224 | 0.144 | 0.668 | | 1 |
| (CVD-I) - (PD-D) | 0.081 | 0.118 | 526 | 0.687 | -0.15 | 0.312 | 0.492 | | 1 |
| (CVD-I) - (PD-MCI) | 0.229 | 0.095 | 526 | 2.419 | 0.043 | 0.415 | **0.016** | | 0.715 |
| (CVD-I) - (PD-CU) | 0.188 | 0.097 | 526 | 1.932 | -0.003 | 0.378 | 0.054 | | 1 |
| (CVD-I) - ALS | -0.367 | 0.105 | 526 | -3.507 | -0.572 | -0.161 | **<0.001** | | **0.022** |

| Contrast | Estimate | SE | df | t-value | 95% CI low | 95% CI high | P-value (raw) | P-value adj. (vs HC)[a] | P-value adj. (vs all pairs)[b] |
|---|---|---|---|---|---|---|---|---|---|
| (CVD-CU) - FTD | -0.249 | 0.09 | 526 | -2.763 | -0.426 | -0.072 | **0.006** | | 0.267 |
| (CVD-CU) - (PD-D) | -0.128 | 0.115 | 526 | -1.109 | -0.354 | 0.099 | 0.268 | | 1 |
| (CVD-CU) - (PD-MCI) | 0.021 | 0.091 | 526 | 0.226 | -0.158 | 0.199 | 0.821 | | 1 |
| (CVD-CU) - (PD-CU) | -0.021 | 0.093 | 526 | -0.228 | -0.203 | 0.161 | 0.820 | | 1 |
| (CVD-CU) - ALS | -0.576 | 0.1 | 526 | -5.757 | -0.772 | -0.379 | **<0.001** | | **<0.001** |
| FTD - (PD-D) | 0.121 | 0.123 | 526 | 0.988 | -0.12 | 0.362 | 0.323 | | 1 |
| FTD - (PD-MCI) | 0.269 | 0.101 | 526 | 2.678 | 0.072 | 0.467 | **0.008** | | 0.344 |
| FTD - (PD-CU) | 0.228 | 0.102 | 526 | 2.224 | 0.027 | 0.429 | **0.027** | | 1 |
| FTD - ALS | -0.327 | 0.109 | 526 | -3.01 | -0.54 | -0.113 | **0.003** | | 0.123 |
| (PD-D) - (PD-MCI) | 0.148 | 0.123 | 526 | 1.202 | -0.094 | 0.391 | 0.230 | | 1 |
| (PD-D) - (PD-CU) | 0.107 | 0.125 | 526 | 0.852 | -0.139 | 0.353 | 0.395 | | 1 |
| (PD-D) - ALS | -0.448 | 0.131 | 526 | -3.423 | -0.705 | -0.191 | **<0.001** | | **0.030** |
| (PD-MCI) - (PD-CU) | -0.042 | 0.103 | 526 | -0.405 | -0.244 | 0.161 | 0.686 | | 1 |
| (PD-MCI) - ALS | -0.596 | 0.11 | 526 | -5.433 | -0.812 | -0.381 | **<0.001** | | **<0.001** |
| (PD-CU) - ALS | -0.554 | 0.11 | 526 | -5.028 | -0.771 | -0.338 | **<0.001** | | **<0.001** |
| **p-tau181** | | | | | | | | | |
| HC - AD | -16.004 | 3.195 | 528 | -5.01 | -22.28 | -9.728 | **<0.001** | **<0.001** | **<0.001** |
| HC - MCI | -6.267 | 2.656 | 528 | -2.359 | -11.485 | -1.049 | **0.019** | 0.168 | 0.840 |
| HC - (CVD-I) | -4.248 | 2.775 | 528 | -1.531 | -9.699 | 1.204 | 0.126 | 1 | 1 |
| HC - (CVD-CU) | -2.752 | 2.653 | 528 | -1.037 | -7.963 | 2.459 | 0.300 | 1 | 1 |
| HC - FTD | -4.449 | 2.905 | 528 | -1.531 | -10.156 | 1.258 | 0.126 | 1 | 1 |
| HC - (PD-D) | -4.042 | 3.535 | 528 | -1.144 | -10.986 | 2.901 | 0.253 | 1 | 1 |
| HC - (PD-MCI) | -2.876 | 2.959 | 528 | -0.972 | -8.688 | 2.936 | 0.331 | 1 | 1 |
| HC - (PD-CU) | -2.864 | 2.985 | 528 | -0.959 | -8.729 | 3.001 | 0.338 | 1 | 1 |
| HC - ALS | -9.359 | 3.123 | 528 | -2.997 | -15.494 | -3.223 | **0.003** | **0.026** | 0.129 |
| AD - MCI | 9.737 | 2.706 | 528 | 3.598 | 4.421 | 15.052 | **<0.001** | | **0.016** |
| AD - (CVD-I) | 11.756 | 2.861 | 528 | 4.109 | 6.135 | 17.377 | **<0.001** | | **0.002** |
| AD - (CVD-CU) | 13.252 | 2.809 | 528 | 4.717 | 7.733 | 18.771 | **<0.001** | | **<0.001** |
| AD - FTD | 11.555 | 2.985 | 528 | 3.87 | 5.69 | 17.42 | **<0.001** | | **0.006** |
| AD - (PD-D) | 11.962 | 3.565 | 528 | 3.356 | 4.959 | 18.965 | **<0.001** | | **0.038** |
| AD - (PD-MCI) | 13.128 | 3.058 | 528 | 4.293 | 7.121 | 19.136 | **<0.001** | | **<0.001** |
| AD - (PD-CU) | 13.14 | 3.13 | 528 | 4.198 | 6.991 | 19.289 | **<0.001** | | **0.001** |

| Contrast | Estimate | SE | df | t-value | 95% CI low | 95% CI high | P-value (raw) | P-value adj. (vs HC)[a] | P-value adj. (vs all pairs)[b] |
|---|---|---|---|---|---|---|---|---|---|
| AD - ALS | 6.645 | 3.285 | 528 | 2.023 | 0.193 | 13.098 | **0.044** | | 1 |
| MCI - (CVD-I) | 2.019 | 2.295 | 528 | 0.88 | -2.489 | 6.528 | 0.379 | | 1 |
| MCI - (CVD-CU) | 3.515 | 2.193 | 528 | 1.603 | -0.794 | 7.824 | 0.110 | | 1 |
| MCI - FTD | 1.818 | 2.465 | 528 | 0.738 | -3.024 | 6.66 | 0.461 | | 1 |
| MCI - (PD-D) | 2.225 | 3.152 | 528 | 0.706 | -3.967 | 8.416 | 0.481 | | 1 |
| MCI - (PD-MCI) | 3.391 | 2.524 | 528 | 1.344 | -1.567 | 8.35 | 0.180 | | 1 |
| MCI - (PD-CU) | 3.403 | 2.591 | 528 | 1.313 | -1.688 | 8.494 | 0.190 | | 1 |
| MCI - ALS | -3.092 | 2.786 | 528 | -1.11 | -8.565 | 2.382 | 0.268 | | 1 |
| (CVD-I) - (CVD-CU) | 1.496 | 2.286 | 528 | 0.654 | -2.994 | 5.986 | 0.513 | | 1 |
| (CVD-I) - FTD | -0.201 | 2.572 | 528 | -0.078 | -5.254 | 4.852 | 0.938 | | 1 |
| (CVD-I) - (PD-D) | 0.206 | 3.229 | 528 | 0.064 | -6.138 | 6.549 | 0.949 | | 1 |
| (CVD-I) - (PD-MCI) | 1.372 | 2.6 | 528 | 0.528 | -3.735 | 6.479 | 0.598 | | 1 |
| (CVD-I) - (PD-CU) | 1.384 | 2.662 | 528 | 0.52 | -3.846 | 6.613 | 0.603 | | 1 |
| (CVD-I) - ALS | -5.111 | 2.869 | 528 | -1.782 | -10.746 | 0.524 | 0.075 | | 1 |
| (CVD-CU) - FTD | -1.697 | 2.46 | 528 | -0.69 | -6.53 | 3.136 | 0.491 | | 1 |
| (CVD-CU) - (PD-D) | -1.29 | 3.149 | 528 | -0.41 | -7.477 | 4.896 | 0.682 | | 1 |
| (CVD-CU) - (PD-MCI) | -0.124 | 2.482 | 528 | -0.05 | -4.999 | 4.751 | 0.960 | | 1 |
| (CVD-CU) - (PD-CU) | -0.112 | 2.531 | 528 | -0.044 | -5.084 | 4.86 | 0.965 | | 1 |
| (CVD-CU) - ALS | -6.607 | 2.73 | 528 | -2.42 | -11.97 | -1.243 | **0.016** | | 0.714 |
| FTD - (PD-D) | 0.407 | 3.353 | 528 | 0.121 | -6.18 | 6.993 | 0.904 | | 1 |
| FTD - (PD-MCI) | 1.573 | 2.754 | 528 | 0.571 | -3.836 | 6.982 | 0.568 | | 1 |
| FTD - (PD-CU) | 1.585 | 2.803 | 528 | 0.566 | -3.921 | 7.091 | 0.572 | | 1 |
| FTD - ALS | -4.91 | 2.97 | 528 | -1.653 | -10.744 | 0.925 | 0.099 | | 1 |
| (PD-D) - (PD-MCI) | 1.166 | 3.376 | 528 | 0.346 | -5.466 | 7.798 | 0.730 | | 1 |
| (PD-D) - (PD-CU) | 1.178 | 3.425 | 528 | 0.344 | -5.551 | 7.907 | 0.731 | | 1 |
| (PD-D) - ALS | -5.316 | 3.58 | 528 | -1.485 | -12.35 | 1.717 | 0.138 | | 1 |
| (PD-MCI) - (PD-CU) | 0.012 | 2.818 | 528 | 0.004 | -5.523 | 5.547 | 0.997 | | 1 |
| (PD-MCI) - ALS | -6.483 | 3.002 | 528 | -2.159 | -12.381 | -0.585 | **0.031** | | 1 |
| (PD-CU) - ALS | -6.495 | 3.017 | 528 | -2.153 | -12.422 | -0.567 | **0.032** | | 1 |
| **Aβ42/40** | | | | | | | | | |
| HC - AD | 2.35e-4 | 0.004 | 508 | 0.067 | -0.007 | 0.007 | 0.947 | 1 | 1 |
| HC - MCI | 0.003 | 0.003 | 508 | 1.127 | -0.002 | 0.009 | 0.260 | 1 | 1 |

| Contrast | Estimate | SE | df | t-value | 95% CI low | 95% CI high | P-value (raw) | P-value adj. (vs HC)[a] | P-value adj. (vs all pairs)[b] |
|---|---|---|---|---|---|---|---|---|---|
| HC - (CVD-I) | -0.001 | 0.003 | 508 | -0.424 | -0.007 | 0.005 | 0.672 | 1 | 1 |
| HC - (CVD-CU) | 3.14e-4 | 0.003 | 508 | 0.11 | -0.005 | 0.006 | 0.913 | 1 | 1 |
| HC - FTD | 0.003 | 0.003 | 508 | 0.808 | -0.004 | 0.009 | 0.419 | 1 | 1 |
| HC - (PD-D) | 3.4e-4 | 0.004 | 508 | 0.089 | -0.007 | 0.008 | 0.929 | 1 | 1 |
| HC - (PD-MCI) | 0.002 | 0.003 | 508 | 0.522 | -0.005 | 0.008 | 0.602 | 1 | 1 |
| HC - (PD-CU) | -0.003 | 0.003 | 508 | -0.946 | -0.009 | 0.003 | 0.344 | 1 | 1 |
| HC - ALS | 0.004 | 0.003 | 508 | 1.094 | -0.003 | 0.01 | 0.275 | 1 | 1 |
| AD - MCI | 0.003 | 0.003 | 508 | 1.005 | -0.003 | 0.009 | 0.316 | | 1 |
| AD - (CVD-I) | -0.001 | 0.003 | 508 | -0.486 | -0.008 | 0.005 | 0.627 | | 1 |
| AD - (CVD-CU) | 7.91e-5 | 0.003 | 508 | 0.026 | -0.006 | 0.006 | 0.980 | | 1 |
| AD - FTD | 0.002 | 0.003 | 508 | 0.702 | -0.004 | 0.009 | 0.483 | | 1 |
| AD - (PD-D) | 1.05e-4 | 0.004 | 508 | 0.027 | -0.007 | 0.008 | 0.978 | | 1 |
| AD - (PD-MCI) | 0.001 | 0.003 | 508 | 0.429 | -0.005 | 0.008 | 0.668 | | 1 |
| AD - (PD-CU) | -0.003 | 0.003 | 508 | -0.959 | -0.01 | 0.003 | 0.338 | | 1 |
| AD - ALS | 0.003 | 0.004 | 508 | 0.96 | -0.004 | 0.01 | 0.337 | | 1 |
| MCI - (CVD-I) | -0.004 | 0.002 | 508 | -1.881 | -0.009 | 1.98e-4 | 0.060 | | 1 |
| MCI - (CVD-CU) | -0.003 | 0.002 | 508 | -1.247 | -0.007 | 0.002 | 0.213 | | 1 |
| MCI - FTD | -6.73e-4 | 0.003 | 508 | -0.255 | -0.006 | 0.005 | 0.799 | | 1 |
| MCI - (PD-D) | -0.003 | 0.003 | 508 | -0.858 | -0.009 | 0.004 | 0.392 | | 1 |
| MCI - (PD-MCI) | -0.002 | 0.003 | 508 | -0.572 | -0.007 | 0.004 | 0.567 | | 1 |
| MCI - (PD-CU) | -0.006 | 0.003 | 508 | -2.281 | -0.012 | -8.67e-4 | **0.023** | | 1 |
| MCI - ALS | 4.49e-4 | 0.003 | 508 | 0.152 | -0.005 | 0.006 | 0.879 | | 1 |
| (CVD-I) - (CVD-CU) | 0.002 | 0.002 | 508 | 0.657 | -0.003 | 0.006 | 0.512 | | 1 |
| (CVD-I) - FTD | 0.004 | 0.003 | 508 | 1.396 | -0.002 | 0.009 | 0.163 | | 1 |
| (CVD-I) - (PD-D) | 0.002 | 0.003 | 508 | 0.47 | -0.005 | 0.008 | 0.639 | | 1 |
| (CVD-I) - (PD-MCI) | 0.003 | 0.003 | 508 | 1.067 | -0.002 | 0.008 | 0.286 | | 1 |
| (CVD-I) - (PD-CU) | -0.002 | 0.003 | 508 | -0.635 | -0.007 | 0.004 | 0.526 | | 1 |
| (CVD-I) - ALS | 0.005 | 0.003 | 508 | 1.63 | -0.001 | 0.011 | 0.104 | | 1 |
| (CVD-CU) - FTD | 0.002 | 0.003 | 508 | 0.842 | -0.003 | 0.007 | 0.400 | | 1 |
| (CVD-CU) - (PD-D) | 2.57e-5 | 0.003 | 508 | 0.008 | -0.007 | 0.007 | 0.994 | | 1 |
| (CVD-CU) - (PD-MCI) | 0.001 | 0.003 | 508 | 0.511 | -0.004 | 0.007 | 0.609 | | 1 |
| (CVD-CU) - (PD-CU) | -0.003 | 0.003 | 508 | -1.248 | -0.009 | 0.002 | 0.212 | | 1 |

| Contrast | Estimate | SE | df | t-value | 95% CI low | 95% CI high | P-value (raw) | P-value adj. (vs HC)[a] | P-value adj. (vs all pairs)[b] |
|---|---|---|---|---|---|---|---|---|---|
| (CVD-CU) - ALS | 0.003 | 0.003 | 508 | 1.158 | -0.002 | 0.009 | 0.247 | | 1 |
| FTD - (PD-D) | -0.002 | 0.004 | 508 | -0.612 | -0.009 | 0.005 | 0.541 | | 1 |
| FTD - (PD-MCI) | -8.71e-4 | 0.003 | 508 | -0.292 | -0.007 | 0.005 | 0.770 | | 1 |
| FTD - (PD-CU) | -0.006 | 0.003 | 508 | -1.856 | -0.011 | 3.25e-4 | 0.064 | | 1 |
| FTD - ALS | 0.001 | 0.003 | 508 | 0.352 | -0.005 | 0.007 | 0.725 | | 1 |
| (PD-D) - (PD-MCI) | 0.001 | 0.004 | 508 | 0.369 | -0.006 | 0.008 | 0.712 | | 1 |
| (PD-D) - (PD-CU) | -0.003 | 0.004 | 508 | -0.925 | -0.011 | 0.004 | 0.355 | | 1 |
| (PD-D) - ALS | 0.003 | 0.004 | 508 | 0.872 | -0.004 | 0.011 | 0.383 | | 1 |
| (PD-MCI) - (PD-CU) | -0.005 | 0.003 | 508 | -1.565 | -0.011 | 0.001 | 0.118 | | 1 |
| (PD-MCI) - ALS | 0.002 | 0.003 | 508 | 0.622 | -0.004 | 0.008 | 0.534 | | 1 |
| (PD-CU) - ALS | 0.007 | 0.003 | 508 | 2.1 | 4.32e-4 | 0.013 | **0.036** | | 1 |
| **NfL** | | | | | | | | | |
| HC - AD | -13.532 | 5.157 | 535 | -2.624 | -23.662 | -3.401 | **0.009** | 0.080 | 0.402 |
| HC - MCI | -7.704 | 4.305 | 535 | -1.79 | -16.161 | 0.753 | 0.074 | 0.667 | 1 |
| HC - (CVD-I) | -12.272 | 4.435 | 535 | -2.767 | -20.985 | -3.559 | **0.006** | 0.053 | 0.264 |
| HC - (CVD-CU) | -9.479 | 4.296 | 535 | -2.207 | -17.917 | -1.041 | **0.028** | 0.250 | 1 |
| HC - FTD | -17.289 | 4.705 | 535 | -3.674 | -26.532 | -8.046 | **<0.001** | **0.002** | **0.012** |
| HC - (PD-D) | -8.605 | 5.713 | 535 | -1.506 | -19.828 | 2.619 | 0.133 | 1 | 1 |
| HC - (PD-MCI) | -3.406 | 4.791 | 535 | -0.711 | -12.817 | 6.005 | 0.477 | 1 | 1 |
| HC - (PD-CU) | -2.102 | 4.835 | 535 | -0.435 | -11.599 | 7.396 | 0.664 | 1 | 1 |
| HC - ALS | -62.997 | 5.056 | 535 | -12.459 | -72.929 | -53.064 | **<0.001** | **<0.001** | **<0.001** |
| AD - MCI | 5.828 | 4.356 | 535 | 1.338 | -2.73 | 14.385 | 0.182 | | 1 |
| AD - (CVD-I) | 1.26 | 4.49 | 535 | 0.281 | -7.56 | 10.079 | 0.779 | | 1 |
| AD - (CVD-CU) | 4.053 | 4.509 | 535 | 0.899 | -4.805 | 12.91 | 0.369 | | 1 |
| AD - FTD | -3.757 | 4.804 | 535 | -0.782 | -13.194 | 5.68 | 0.435 | | 1 |
| AD - (PD-D) | 4.927 | 5.74 | 535 | 0.858 | -6.348 | 16.202 | 0.391 | | 1 |
| AD - (PD-MCI) | 10.126 | 4.918 | 535 | 2.059 | 0.465 | 19.786 | **0.040** | | 1 |
| AD - (PD-CU) | 11.43 | 5.031 | 535 | 2.272 | 1.547 | 21.313 | **0.023** | | 1 |
| AD - ALS | -49.465 | 5.28 | 535 | -9.368 | -59.837 | -39.093 | **<0.001** | | **<0.001** |
| MCI - (CVD-I) | -4.568 | 3.59 | 535 | -1.272 | -11.62 | 2.485 | 0.204 | | 1 |
| MCI - (CVD-CU) | -1.775 | 3.523 | 535 | -0.504 | -8.696 | 5.146 | 0.615 | | 1 |
| MCI - FTD | -9.585 | 3.971 | 535 | -2.414 | -17.385 | -1.784 | **0.016** | | 0.726 |

| Contrast | Estimate | SE | df | t-value | 95% CI low | 95% CI high | P-value (raw) | P-value adj. (vs HC)[a] | P-value adj. (vs all pairs)[b] |
|---|---|---|---|---|---|---|---|---|---|
| MCI - (PD-D) | -0.901 | 5.078 | 535 | -0.177 | -10.877 | 9.075 | 0.859 | | 1 |
| MCI - (PD-MCI) | 4.298 | 4.065 | 535 | 1.057 | -3.687 | 12.282 | 0.291 | | 1 |
| MCI - (PD-CU) | 5.602 | 4.172 | 535 | 1.343 | -2.593 | 13.797 | 0.180 | | 1 |
| MCI - ALS | -55.293 | 4.485 | 535 | -12.327 | -64.104 | -46.482 | **<0.001** | | **<0.001** |
| (CVD-I) - (CVD-CU) | 2.793 | 3.598 | 535 | 0.776 | -4.275 | 9.861 | 0.438 | | 1 |
| (CVD-I) - FTD | -5.017 | 4.063 | 535 | -1.235 | -12.999 | 2.965 | 0.217 | | 1 |
| (CVD-I) - (PD-D) | 3.667 | 5.134 | 535 | 0.714 | -6.419 | 13.753 | 0.475 | | 1 |
| (CVD-I) - (PD-MCI) | 8.866 | 4.117 | 535 | 2.153 | 0.777 | 16.954 | **0.032** | | 1 |
| (CVD-I) - (PD-CU) | 10.17 | 4.228 | 535 | 2.405 | 1.864 | 18.476 | **0.017** | | 0.743 |
| (CVD-I) - ALS | -50.725 | 4.568 | 535 | -11.104 | -59.699 | -41.751 | **<0.001** | | **<0.001** |
| (CVD-CU) - FTD | -7.809 | 3.955 | 535 | -1.975 | -15.578 | -0.041 | **0.049** | | 1 |
| (CVD-CU) - (PD-D) | 0.874 | 5.067 | 535 | 0.173 | -9.079 | 10.828 | 0.863 | | 1 |
| (CVD-CU) - (PD-MCI) | 6.073 | 3.99 | 535 | 1.522 | -1.764 | 13.91 | 0.129 | | 1 |
| (CVD-CU) - (PD-CU) | 7.377 | 4.069 | 535 | 1.813 | -0.615 | 15.37 | 0.070 | | 1 |
| (CVD-CU) - ALS | -53.518 | 4.39 | 535 | -12.191 | -62.142 | -44.894 | **<0.001** | | **<0.001** |
| FTD - (PD-D) | 8.684 | 5.403 | 535 | 1.607 | -1.929 | 19.297 | 0.109 | | 1 |
| FTD - (PD-MCI) | 13.882 | 4.437 | 535 | 3.129 | 5.167 | 22.598 | **0.002** | | 0.083 |
| FTD - (PD-CU) | 15.187 | 4.515 | 535 | 3.364 | 6.318 | 24.056 | **<0.001** | | **0.037** |
| FTD - ALS | -45.708 | 4.784 | 535 | -9.554 | -55.106 | -36.31 | **<0.001** | | **<0.001** |
| (PD-D) - (PD-MCI) | 5.199 | 5.439 | 535 | 0.956 | -5.487 | 15.884 | 0.340 | | 1 |
| (PD-D) - (PD-CU) | 6.503 | 5.518 | 535 | 1.179 | -4.336 | 17.342 | 0.239 | | 1 |
| (PD-D) - ALS | -54.392 | 5.767 | 535 | -9.432 | -65.721 | -43.063 | **<0.001** | | **<0.001** |
| (PD-MCI) - (PD-CU) | 1.304 | 4.54 | 535 | 0.287 | -7.614 | 10.223 | 0.774 | | 1 |
| (PD-MCI) - ALS | -59.591 | 4.837 | 535 | -12.32 | -69.092 | -50.089 | **<0.001** | | **<0.001** |
| (PD-CU) - ALS | -60.895 | 4.862 | 535 | -12.526 | -70.445 | -51.345 | **<0.001** | | **<0.001** |
| **GFAP** | | | | | | | | | |
| HC - AD | -92.272 | 17.729 | 535 | -5.205 | -127.099 | -57.444 | **<0.001** | **<0.001** | **<0.001** |
| HC - MCI | -32.45 | 14.8 | 535 | -2.193 | -61.523 | -3.377 | **0.029** | 0.259 | 1 |
| HC - (CVD-I) | -18.531 | 15.248 | 535 | -1.215 | -48.485 | 11.422 | 0.225 | 1 | 1 |
| HC - (CVD-CU) | -18.301 | 14.768 | 535 | -1.239 | -47.311 | 10.709 | 0.216 | 1 | 1 |
| HC - FTD | -31.464 | 16.176 | 535 | -1.945 | -63.24 | 0.311 | 0.052 | 0.471 | 1 |
| HC - (PD-D) | -26.725 | 19.642 | 535 | -1.361 | -65.309 | 11.859 | 0.174 | 1 | 1 |

| Contrast | Estimate | SE | df | t-value | 95% CI low | 95% CI high | P-value (raw) | P-value adj. (vs HC)[a] | P-value adj. (vs all pairs)[b] |
|---|---|---|---|---|---|---|---|---|---|
| HC - (PD-MCI) | -13.519 | 16.47 | 535 | -0.821 | -45.873 | 18.835 | 0.412 | 1 | 1 |
| HC - (PD-CU) | -2.802 | 16.621 | 535 | -0.169 | -35.453 | 29.85 | 0.866 | 1 | 1 |
| HC - ALS | -33.687 | 17.382 | 535 | -1.938 | -67.833 | 0.459 | 0.053 | 0.478 | 1 |
| AD - MCI | 59.822 | 14.976 | 535 | 3.995 | 30.403 | 89.241 | **<0.001** | | **0.003** |
| AD - (CVD-I) | 73.74 | 15.435 | 535 | 4.778 | 43.42 | 104.06 | **<0.001** | | **<0.001** |
| AD - (CVD-CU) | 73.971 | 15.502 | 535 | 4.772 | 43.52 | 104.423 | **<0.001** | | **<0.001** |
| AD - FTD | 60.807 | 16.516 | 535 | 3.682 | 28.364 | 93.251 | **<0.001** | | **0.011** |
| AD - (PD-D) | 65.547 | 19.732 | 535 | 3.322 | 26.785 | 104.308 | **<0.001** | | **0.043** |
| AD - (PD-MCI) | 78.752 | 16.907 | 535 | 4.658 | 45.541 | 111.964 | **<0.001** | | **<0.001** |
| AD - (PD-CU) | 89.47 | 17.296 | 535 | 5.173 | 55.493 | 123.447 | **<0.001** | | **<0.001** |
| AD - ALS | 58.585 | 18.152 | 535 | 3.228 | 22.928 | 94.242 | **0.001** | | 0.060 |
| MCI - (CVD-I) | 13.918 | 12.342 | 535 | 1.128 | -10.327 | 38.163 | 0.260 | | 1 |
| MCI - (CVD-CU) | 14.149 | 12.112 | 535 | 1.168 | -9.643 | 37.941 | 0.243 | | 1 |
| MCI - FTD | 0.985 | 13.652 | 535 | 0.072 | -25.833 | 27.803 | 0.943 | | 1 |
| MCI - (PD-D) | 5.725 | 17.458 | 535 | 0.328 | -28.57 | 40.019 | 0.743 | | 1 |
| MCI - (PD-MCI) | 18.93 | 13.974 | 535 | 1.355 | -8.52 | 46.38 | 0.176 | | 1 |
| MCI - (PD-CU) | 29.648 | 14.342 | 535 | 2.067 | 1.475 | 57.821 | **0.039** | | 1 |
| MCI - ALS | -1.237 | 15.42 | 535 | -0.08 | -31.529 | 29.054 | 0.936 | | 1 |
| (CVD-I) - (CVD-CU) | 0.231 | 12.37 | 535 | 0.019 | -24.068 | 24.53 | 0.985 | | 1 |
| (CVD-I) - FTD | -12.933 | 13.969 | 535 | -0.926 | -40.373 | 14.507 | 0.355 | | 1 |
| (CVD-I) - (PD-D) | -8.194 | 17.651 | 535 | -0.464 | -42.867 | 26.48 | 0.643 | | 1 |
| (CVD-I) - (PD-MCI) | 5.012 | 14.155 | 535 | 0.354 | -22.794 | 32.819 | 0.723 | | 1 |
| (CVD-I) - (PD-CU) | 15.73 | 14.536 | 535 | 1.082 | -12.825 | 44.284 | 0.280 | | 1 |
| (CVD-I) - ALS | -15.155 | 15.705 | 535 | -0.965 | -46.007 | 15.696 | 0.335 | | 1 |
| (CVD-CU) - FTD | -13.164 | 13.596 | 535 | -0.968 | -39.872 | 13.544 | 0.333 | | 1 |
| (CVD-CU) - (PD-D) | -8.424 | 17.419 | 535 | -0.484 | -42.642 | 25.793 | 0.629 | | 1 |
| (CVD-CU) - (PD-MCI) | 4.781 | 13.716 | 535 | 0.349 | -22.162 | 31.725 | 0.728 | | 1 |
| (CVD-CU) - (PD-CU) | 15.499 | 13.987 | 535 | 1.108 | -11.978 | 42.976 | 0.268 | | 1 |
| (CVD-CU) - ALS | -15.386 | 15.092 | 535 | -1.019 | -45.034 | 14.261 | 0.308 | | 1 |
| FTD - (PD-D) | 4.739 | 18.574 | 535 | 0.255 | -31.747 | 41.226 | 0.799 | | 1 |
| FTD - (PD-MCI) | 17.945 | 15.252 | 535 | 1.177 | -12.016 | 47.907 | 0.240 | | 1 |
| FTD - (PD-CU) | 28.663 | 15.521 | 535 | 1.847 | -1.827 | 59.152 | 0.065 | | 1 |

| Contrast | Estimate | SE | df | t-value | 95% CI low | 95% CI high | P-value (raw) | P-value adj. (vs HC)[a] | P-value adj. (vs all pairs)[b] |
|---|---|---|---|---|---|---|---|---|---|
| FTD - ALS | -2.222 | 16.447 | 535 | -0.135 | -34.531 | 30.087 | 0.893 | | 1 |
| (PD-D) - (PD-MCI) | 13.206 | 18.7 | 535 | 0.706 | -23.528 | 49.94 | 0.480 | | 1 |
| (PD-D) - (PD-CU) | 23.923 | 18.969 | 535 | 1.261 | -13.34 | 61.187 | 0.208 | | 1 |
| (PD-D) - ALS | -6.962 | 19.826 | 535 | -0.351 | -45.908 | 31.985 | 0.726 | | 1 |
| (PD-MCI) - (PD-CU) | 10.718 | 15.608 | 535 | 0.687 | -19.943 | 41.378 | 0.493 | | 1 |
| (PD-MCI) - ALS | -20.167 | 16.628 | 535 | -1.213 | -52.832 | 12.497 | 0.226 | | 1 |
| (PD-CU) - ALS | -30.885 | 16.713 | 535 | -1.848 | -63.717 | 1.947 | 0.065 | | 1 |

Estimated marginal means pairwise contrasts. *APOE* = apolipoprotein E; Aβ42/40 = amyloid-β42 to amyloid-β40 ratio; CI = confidence interval; df = degrees of freedom; SE = standard error; GFAP = glial fibrillary acidic protein; NfL = neurofilament light chain; p-tau181 = tau phosphorylated at threonine 181; p-tau217 = tau phosphorylated at threonine 217; AD = Alzheimer's disease; ALS = amyotrophic lateral sclerosis; CVD-CU = cerebrovascular disease, cognitively unimpaired; CVD-I = cerebrovascular disease, cognitively impaired; FTD = frontotemporal dementia; HC = healthy controls; MCI = mild cognitive impairment; PD-CU = Parkinson's disease, cognitively unimpaired; PD-D = Parkinson's disease with dementia; PD-MCI = Parkinson's disease with mild cognitive impairment.

[a] P-values Bonferroni-adjusted across nine comparisons with HC per biomarker; blank for comparisons not involving HC.

[b] P-values Bonferroni-adjusted across 45 pairwise comparisons per biomarker.

### Inferred amyloid-β positivity

***Table S3.*** *Amyloid-β positivity classification by diagnostic group using published p-tau217 reference thresholds.*

| Inferred Aβ status | HC | MCI | AD | PD-CU | PD-MCI | PD-D | FTD | CVD-CU | CVD-I | ALS | Statistic[a] |
|---|---|---|---|---|---|---|---|---|---|---|---|
| Positive (>0.63 pg/mL) | 4 (9.3) | 34 (40) | 33 (82.5) | 4 (8.3) | 3 (5.9) | 8 (30.8) | 15 (28.3) | 3 (3.6) | 21 (30.4) | 22 (55) | $\chi 2(18) = 142.3$, **$P < 0.001$** |
| Intermediate | 11 (25.6) | 16 (18.8) | 4 (10) | 9 (18.8) | 12 (23.5) | 2 (7.7) | 10 (18.9) | 21 (25) | 13 (18.8) | 8 (20) | |
| Negative (<0.40 pg/mL) | 28 (65.1) | 35 (41.2) | 3 (7.5) | 35 (72.9) | 36 (70.6) | 16 (61.5) | 28 (52.8) | 60 (71.4) | 35 (50.7) | 10 (25) | |

Values are n (%). Plasma p-tau217 reference points from Ashton et al., 2024. Negative: <0.40 pg/mL (95% sensitivity). Intermediate: 0.40–0.63 pg/mL. Positive: >0.63 pg/mL (95% specificity). Aβ = amyloid-β; p-tau217 = tau phosphorylated at threonine 217; AD = Alzheimer's disease; ALS = amyotrophic lateral sclerosis; CVD-CU = cerebrovascular disease, cognitively unimpaired; CVD-I = cerebrovascular disease, cognitively impaired; FTD = frontotemporal dementia; HC = healthy controls; MCI = mild cognitive impairment; PD-CU = Parkinson's disease, cognitively unimpaired; PD-D = Parkinson's disease with dementia; PD-MCI = Parkinson's disease with mild cognitive impairment.

[a] Overall Pearson chi-square test comparing the three-category inferred Aβ status distributions across diagnostic groups. The result applies to the whole table, not only the positive-status row.

***Table S4.*** *All pairwise Dunn's test comparisons for inferred amyloid-β positivity classification across diagnostic groups.*

| Group 1 | Group 2 | N1 | N2 | Z | P-value (raw) | P-value adj. (vs HC)[a] | P-value adj. (vs all pairs)[b] |
|---|---|---|---|---|---|---|---|
| HC | MCI | 43 | 85 | 3.244 | **0.001** | **0.011** | 0.053 |
| HC | AD | 43 | 40 | 6.623 | **<0.001** | **<0.001** | **<0.001** |
| HC | PD-CU | 43 | 48 | -0.557 | 0.578 | 1 | 1 |
| HC | PD-MCI | 43 | 51 | -0.515 | 0.607 | 1 | 1 |
| HC | PD-D | 43 | 26 | 0.974 | 0.330 | 1 | 1 |
| HC | FTD | 43 | 53 | 1.66 | 0.097 | 0.873 | 1 |
| HC | CVD-CU | 43 | 84 | -0.741 | 0.459 | 1 | 1 |
| HC | CVD-I | 43 | 69 | 2.002 | **0.045** | 0.408 | 1 |
| HC | ALS | 43 | 40 | 4.397 | **<0.001** | **<0.001** | **<0.001** |
| MCI | AD | 85 | 40 | 4.422 | **<0.001** | | **<0.001** |
| MCI | PD-CU | 85 | 48 | -4.01 | **<0.001** | | **0.003** |
| MCI | PD-MCI | 85 | 51 | -4.029 | **<0.001** | | **0.003** |
| MCI | PD-D | 85 | 26 | -1.629 | 0.103 | | 1 |
| MCI | FTD | 85 | 53 | -1.522 | 0.128 | | 1 |
| MCI | CVD-CU | 85 | 84 | -4.849 | **<0.001** | | **<0.001** |
| MCI | CVD-I | 85 | 69 | -1.346 | 0.178 | | 1 |
| MCI | ALS | 85 | 40 | 1.871 | 0.061 | | 1 |
| AD | PD-CU | 40 | 48 | -7.342 | **<0.001** | | **<0.001** |
| AD | PD-MCI | 40 | 51 | -7.393 | **<0.001** | | **<0.001** |
| AD | PD-D | 40 | 26 | -4.815 | **<0.001** | | **<0.001** |
| AD | FTD | 40 | 53 | -5.32 | **<0.001** | | **<0.001** |
| AD | CVD-CU | 40 | 84 | -8.296 | **<0.001** | | **<0.001** |
| AD | CVD-I | 40 | 69 | -5.364 | **<0.001** | | **<0.001** |
| AD | ALS | 40 | 40 | -2.187 | **0.029** | | 1 |
| PD-CU | PD-MCI | 48 | 51 | 0.052 | 0.959 | | 1 |
| PD-CU | PD-D | 48 | 26 | 1.473 | 0.141 | | 1 |
| PD-CU | FTD | 48 | 53 | 2.296 | **0.022** | | 0.975 |
| PD-CU | CVD-CU | 48 | 84 | -0.122 | 0.903 | | 1 |
| PD-CU | CVD-I | 48 | 69 | 2.691 | **0.007** | | 0.320 |
| PD-CU | ALS | 48 | 40 | 5.057 | **<0.001** | | **<0.001** |
| PD-MCI | PD-D | 51 | 26 | 1.446 | 0.148 | | 1 |
| PD-MCI | FTD | 51 | 53 | 2.28 | **0.023** | | 1 |
| PD-MCI | CVD-CU | 51 | 84 | -0.182 | 0.855 | | 1 |
| PD-MCI | CVD-I | 51 | 69 | 2.683 | **0.007** | | 0.328 |
| PD-MCI | ALS | 51 | 40 | 5.077 | **<0.001** | | **<0.001** |
| PD-D | FTD | 26 | 53 | 0.412 | 0.680 | | 1 |
| PD-D | CVD-CU | 26 | 84 | -1.697 | 0.090 | | 1 |
| PD-D | CVD-I | 26 | 69 | 0.639 | 0.523 | | 1 |
| PD-D | ALS | 26 | 40 | 2.874 | **0.004** | | 0.183 |
| FTD | CVD-CU | 53 | 84 | -2.733 | **0.006** | | 0.282 |
| FTD | CVD-I | 53 | 69 | 0.265 | 0.791 | | 1 |
| FTD | ALS | 53 | 40 | 2.985 | **0.003** | | 0.128 |
| CVD-CU | CVD-I | 84 | 69 | 3.249 | **0.001** | | 0.052 |
| CVD-CU | ALS | 84 | 40 | 5.751 | **<0.001** | | **<0.001** |

| Group 1 | Group 2 | N1 | N2 | Z | P-value (raw) | P-value adj. (vs HC)[a] | P-value adj. (vs all pairs)[b] |
|---|---|---|---|---|---|---|---|
| CVD-I | ALS | 69 | 40 | 2.903 | **0.004** | | 0.166 |

Dunn's Z-statistic with rank-biserial correlation effect size. Overall Kruskal-Wallis test: $\chi^2(9) = 116.22$, **P < 0.001**. AD = Alzheimer's disease; ALS = amyotrophic lateral sclerosis; CVD-CU = cerebrovascular disease, cognitively unimpaired; CVD-I = cerebrovascular disease, cognitively impaired; FTD = frontotemporal dementia; HC = healthy controls; MCI = mild cognitive impairment; PD-CU = Parkinson's disease, cognitively unimpaired; PD-D = Parkinson's disease with dementia; PD-MCI = Parkinson's disease with mild cognitive impairment.

[a] P-values Bonferroni-adjusted across nine comparisons with HC; blank for comparisons not involving HC.

[b] P-values Bonferroni-adjusted across 45 pairwise comparisons.

***Table S5.*** *Binomial risk differences for inferred amyloid-β positivity relative to healthy controls, using the lower (p-tau217 > 0.40 pg/mL) and upper (p-tau217 > 0.63 pg/mL) cut-off thresholds from Ashton et al., 2024.*

| **Diagnostic Group** | **Risk Difference vs HC** | **Standard Error** | **95% CI Lower** | **95% CI Upper** | **Z** | **P-value (raw)** | **P-value adj. (vs HC)[a]** |
|---|---|---|---|---|---|---|---|
| Lower cut-off (>0.40 pg/mL) | | | | | | | |
| MCI | 0.239 | 0.09 | 0.063 | 0.416 | 2.655 | **0.008** | 0.071 |
| AD | 0.576 | 0.084 | 0.412 | 0.74 | 6.878 | **<0.001** | **<0.001** |
| PD-CU | -0.078 | 0.097 | -0.268 | 0.112 | -0.805 | 0.421 | 1 |
| PD-MCI | -0.055 | 0.097 | -0.244 | 0.135 | -0.566 | 0.572 | 1 |
| PD-D | 0.036 | 0.12 | -0.199 | 0.271 | 0.298 | 0.765 | 1 |
| FTD | 0.123 | 0.1 | -0.073 | 0.319 | 1.23 | 0.219 | 1 |
| CVD-CU | -0.063 | 0.088 | -0.235 | 0.109 | -0.719 | 0.472 | 1 |
| CVD-I | 0.144 | 0.094 | -0.041 | 0.329 | 1.525 | 0.127 | 1 |
| ALS | 0.401 | 0.1 | 0.205 | 0.597 | 4.018 | **<0.001** | **<0.001** |
| Upper cut-off (>0.63 pg/mL) | | | | | | | |
| MCI | 0.307 | 0.069 | 0.171 | 0.443 | 4.437 | **<0.001** | **<0.001** |
| AD | 0.732 | 0.075 | 0.586 | 0.878 | 9.806 | **<0.001** | **<0.001** |
| PD-CU | -0.01 | 0.06 | -0.127 | 0.107 | -0.163 | 0.871 | 1 |
| PD-MCI | -0.034 | 0.055 | -0.142 | 0.074 | -0.62 | 0.536 | 1 |
| PD-D | 0.215 | 0.101 | 0.017 | 0.412 | 2.13 | **0.033** | 0.298 |
| FTD | 0.19 | 0.076 | 0.041 | 0.339 | 2.497 | **0.013** | 0.113 |
| CVD-CU | -0.057 | 0.049 | -0.153 | 0.038 | -1.177 | 0.239 | 1 |
| CVD-I | 0.211 | 0.071 | 0.072 | 0.35 | 2.98 | **0.003** | **0.026** |
| ALS | 0.457 | 0.09 | 0.28 | 0.634 | 5.062 | **<0.001** | **<0.001** |

Risk difference: absolute difference in proportion classified as positive vs. HC. Lower cut-off threshold: p-tau217 > 0.40 pg/mL. Upper cut-off threshold: p-tau217 > 0.63 pg/mL. CI = confidence interval; p-tau217 = tau phosphorylated at threonine 217; AD = Alzheimer's disease; ALS = amyotrophic lateral sclerosis; CVD-CU = cerebrovascular disease, cognitively unimpaired; CVD-I = cerebrovascular disease, cognitively impaired; FTD = frontotemporal dementia; HC = healthy controls; MCI = mild cognitive impairment; PD-CU = Parkinson's disease, cognitively unimpaired; PD-D = Parkinson's disease with dementia; PD-MCI = Parkinson's disease with mild cognitive impairment.

[a] P-values Bonferroni-adjusted across nine comparisons with HC, separately for each threshold.

#### ALS biomarker-clinical associations

***Table S6.*** *Baseline cross-sectional associations between plasma biomarkers and ALS clinical features. Ordinary least-squares regression adjusted for age, sex, and APOE ε4 allele dosage.*

| Biomarker | Estimate | 95% CI low | 95% CI high | SE | Statistic[a] | df | Partial $R^2$ | P-value (raw) | P-value adj. (Bonferroni)[b] |
|---|---|---|---|---|---|---|---|---|---|
| Upper motor neuron burden | | | | | | | | | |
| p-tau217 | 0.181 | -0.213 | 0.575 | 0.194 | 0.932 | 35 | 0.024 | 0.358 | 1 |
| p-tau181 | -0.014 | -0.046 | 0.019 | 0.016 | -0.842 | 35 | 0.02 | 0.405 | 1 |
| Aβ42/40 | -13.082 | -31.168 | 5.005 | 8.9 | -1.47 | 34 | 0.06 | 0.151 | 0.754 |
| NfL | 0.006 | -5.91e-5 | 0.011 | 0.003 | 2.009 | 35 | 0.103 | 0.052 | 0.262 |
| GFAP | -0.001 | -0.005 | 0.003 | 0.002 | -0.595 | 35 | 0.01 | 0.556 | 1 |
| Lower motor neuron burden | | | | | | | | | |
| p-tau217 | 0.395 | -0.128 | 0.919 | 0.258 | 1.533 | 35 | 0.063 | 0.134 | 0.672 |
| p-tau181 | -0.006 | -0.05 | 0.039 | 0.022 | -0.267 | 35 | 0.002 | 0.791 | 1 |
| Aβ42/40 | 14.466 | -10.348 | 39.28 | 12.21 | 1.185 | 34 | 0.04 | 0.244 | 1 |
| NfL | 0.009 | 0.001 | 0.016 | 0.004 | 2.378 | 35 | 0.139 | **0.023** | 0.115 |
| GFAP | 7.68e-4 | -0.004 | 0.006 | 0.003 | 0.298 | 35 | 0.003 | 0.767 | 1 |
| ALS duration (months) | | | | | | | | | |
| p-tau217 | 5.263 | -2.205 | 12.732 | 3.679 | 1.431 | 35 | 0.055 | 0.161 | 0.807 |
| p-tau181 | -0.24 | -0.869 | 0.389 | 0.31 | -0.773 | 35 | 0.017 | 0.444 | 1 |
| Aβ42/40 | 70.12 | -297.343 | 437.582 | 180.816 | 0.388 | 34 | 0.004 | 0.701 | 1 |
| NfL | -0.1 | -0.209 | 0.008 | 0.054 | -1.872 | 35 | 0.091 | 0.070 | 0.348 |
| GFAP | 0.034 | -0.04 | 0.107 | 0.036 | 0.93 | 35 | 0.024 | 0.359 | 1 |

Estimate = regression coefficient (β); Partial $R^2$ = proportion of variance explained by biomarker after adjusting for covariates. ALS = amyotrophic lateral sclerosis; *APOE* = apolipoprotein E; Aβ42/40 = amyloid-β42 to amyloid-β40 ratio; CI = confidence interval; df = degrees of freedom; SE = standard error; GFAP = glial fibrillary acidic protein; NfL = neurofilament light chain; p-tau181 = tau phosphorylated at threonine 181; p-tau217 = tau phosphorylated at threonine 217.

[a] Regression-coefficient t statistic.

[b] P-values Bonferroni-adjusted across five biomarkers per outcome.

***Table S7.*** *Longitudinal associations between baseline plasma biomarkers and ALSFRS-R decline from the linear mixed-effects model: ALSFRS-R ~ Time × Biomarker + Age + Sex + APOE ε4 + (1 + Time | Subject).*

| **Term** | **Estimate** | **SE** | **Statistic**[a] | **df** | **P-value (raw)** | **P-value adj. (Bonferroni)**[b] |
|---|---|---|---|---|---|---|
| p-tau217 | | | | | | |
| p-tau217 | -1.654 | 1.021 | -1.62 | 33.405 | 0.115 | 0.573 |
| Time × p-tau217 | 1.312 | 1.721 | 0.762 | 16.544 | 0.457 | 1 |
| p-tau181 | | | | | | |
| p-tau181 | -0.03 | 0.092 | -0.324 | 41.248 | 0.747 | 1 |
| Time × p-tau181 | -0.087 | 0.181 | -0.481 | 29.615 | 0.634 | 1 |
| NfL | | | | | | |
| NfL | -0.005 | 0.017 | -0.28 | 43.734 | 0.781 | 1 |
| Time × NfL | -0.096 | 0.037 | -2.587 | 36.753 | **0.014** | 0.069 |
| GFAP | | | | | | |
| GFAP | 0.006 | 0.01 | 0.549 | 34.173 | 0.586 | 1 |
| Time × GFAP | 0.021 | 0.015 | 1.401 | 17.939 | 0.178 | 0.892 |

Time × Biomarker interaction tests whether baseline biomarker levels modify the rate of ALSFRS-R decline. ALS = amyotrophic lateral sclerosis; ALSFRS-R = Amyotrophic Lateral Sclerosis Functional Rating Scale-Revised; *APOE* = apolipoprotein E; df = degrees of freedom; SE = standard error; GFAP = glial fibrillary acidic protein; NfL = neurofilament light chain; p-tau181 = tau phosphorylated at threonine 181; p-tau217 = tau phosphorylated at threonine 217.

[a] Mixed-effects model fixed-effect t statistic

[b] P-values Bonferroni-adjusted across five biomarkers.

***Table S8.*** *Longitudinal associations between baseline plasma biomarkers and MoCA change from the linear mixed-effects model: MoCA ~ Time × Biomarker + Age + Education + Sex + APOE ε4 + (1 + Time | Subject).*

| **Term** | **Estimate** | **SE** | **Statistic**[a] | **df** | **P-value (raw)** | **P-value adj. (Bonferroni)**[b] |
|---|---|---|---|---|---|---|
| p-tau217 | | | | | | |
| p-tau217 | 0.7 | 0.621 | 1.127 | 34.546 | 0.267 | 1 |
| Time × p-tau217 | 0.01 | 0.434 | 0.023 | 3.25 | 0.983 | 1 |
| p-tau181 | | | | | | |
| p-tau181 | 0.031 | 0.057 | 0.543 | 51.159 | 0.589 | 1 |
| Time × p-tau181 | -0.085 | 0.072 | -1.177 | 19.014 | 0.254 | 1 |
| NfL | | | | | | |
| NfL | 0.012 | 0.01 | 1.116 | 54.799 | 0.269 | 1 |
| Time × NfL | -0.026 | 0.017 | -1.509 | 19.429 | 0.147 | 0.737 |
| GFAP | | | | | | |
| GFAP | -0.002 | 0.006 | -0.323 | 42.998 | 0.749 | 1 |
| Time × GFAP | 0.003 | 0.005 | 0.644 | 11.145 | 0.532 | 1 |

Time × Biomarker interaction tests whether baseline biomarker levels modify the rate of MoCA change. ALS = amyotrophic lateral sclerosis; *APOE* = apolipoprotein E; df = degrees of freedom; MoCA = Montreal Cognitive Assessment; SE = standard error; GFAP = glial fibrillary acidic protein; NfL = neurofilament light chain; p-tau181 = tau phosphorylated at threonine 181; p-tau217 = tau phosphorylated at threonine 217.

[a] Mixed-effects model fixed-effect t statistic

[b] P-values Bonferroni-adjusted across five biomarkers.

***Table S9.*** *Longitudinal associations between baseline plasma biomarkers and neuropsychological domain z-scores from the linear mixed-effects model: Domain z-score ~ Time × Biomarker + Age + Education + Sex + APOE ε4 + (1 + Time | Subject).*

| Biomarker | Term | Estimate | SE | Statistic [a] | df | P-value (raw) | P-value adj. (Bonferroni)[b] |
|---|---|---|---|---|---|---|---|
| **Attention & Working Memory** | | | | | | | |
| p-tau181 | p-tau181 | 5.04e-4 | 0.015 | 0.034 | 37.557 | 0.973 | 1 |
| p-tau181 | Time × p-tau181 | -0.002 | 0.008 | -0.272 | 77.47 | 0.787 | 1 |
| p-tau217 | p-tau217 | 0.229 | 0.173 | 1.324 | 34.87 | 0.194 | 0.971 |
| p-tau217 | Time × p-tau217 | -0.017 | 0.04 | -0.428 | 75.026 | 0.670 | 1 |
| NfL | NfL | 0.001 | 0.003 | 0.419 | 38.349 | 0.677 | 1 |
| NfL | Time × NfL | 0.002 | 0.002 | 0.895 | 77.925 | 0.373 | 1 |
| GFAP | GFAP | 0.001 | 0.002 | 0.616 | 35.491 | 0.542 | 1 |
| GFAP | Time × GFAP | -1.23e-4 | 4.59e-4 | -0.269 | 75.584 | 0.789 | 1 |
| Aβ42/40 | Aβ42/40 | 8.49 | 8.61 | 0.986 | 36.323 | 0.331 | 1 |
| Aβ42/40 | Time × Aβ42/40 | 2.282 | 3.561 | 0.641 | 75.266 | 0.523 | 1 |
| **Executive Function** | | | | | | | |
| p-tau181 | p-tau181 | -0.012 | 0.012 | -1.016 | 38.089 | 0.316 | 1 |
| p-tau181 | Time × p-tau181 | 0.001 | 0.007 | 0.208 | 78.056 | 0.836 | 1 |
| p-tau217 | p-tau217 | 0.037 | 0.14 | 0.262 | 35.344 | 0.795 | 1 |
| p-tau217 | Time × p-tau217 | 0.031 | 0.032 | 0.953 | 75.542 | 0.344 | 1 |
| NfL | NfL | -0.001 | 0.002 | -0.619 | 38.641 | 0.540 | 1 |
| NfL | Time × NfL | 0.001 | 0.001 | 0.96 | 78.228 | 0.340 | 1 |
| GFAP | GFAP | 4.53e-4 | 0.001 | 0.33 | 35.551 | 0.743 | 1 |
| GFAP | Time × GFAP | -2.21e-4 | 3.74e-4 | -0.592 | 75.731 | 0.556 | 1 |
| Aβ42/40 | Aβ42/40 | 4.039 | 6.952 | 0.581 | 36.62 | 0.565 | 1 |
| Aβ42/40 | Time × Aβ42/40 | 0.675 | 2.971 | 0.227 | 75.771 | 0.821 | 1 |
| **Language** | | | | | | | |
| p-tau181 | p-tau181 | -0.008 | 0.014 | -0.607 | 36.606 | 0.548 | 1 |
| p-tau181 | Time × p-tau181 | -0.006 | 0.007 | -0.856 | 71.554 | 0.395 | 1 |
| p-tau217 | p-tau217 | 0.129 | 0.163 | 0.791 | 34.711 | 0.434 | 1 |
| p-tau217 | Time × p-tau217 | 0.025 | 0.033 | 0.761 | 69.837 | 0.450 | 1 |
| NfL | NfL | 0.002 | 0.003 | 0.827 | 37.416 | 0.414 | 1 |
| NfL | Time × NfL | -1.22e-4 | 0.002 | -0.074 | 72.282 | 0.941 | 1 |
| GFAP | GFAP | 4.83e-4 | 0.002 | 0.302 | 34.945 | 0.765 | 1 |
| GFAP | Time × GFAP | 4.87e-4 | 3.82e-4 | 1.276 | 70.064 | 0.206 | 1 |
| Aβ42/40 | Aβ42/40 | 3.937 | 8.148 | 0.483 | 35.275 | 0.632 | 1 |
| Aβ42/40 | Time × Aβ42/40 | 4.844 | 2.991 | 1.619 | 69.172 | 0.110 | 0.550 |
| **Verbal Memory** | | | | | | | |
| p-tau181 | p-tau181 | 0.002 | 0.015 | 0.127 | 37.297 | 0.900 | 1 |
| p-tau181 | Time × p-tau181 | 0.013 | 0.009 | 1.377 | 74.112 | 0.173 | 0.864 |

| Biomarker | Term | Estimate | SE | Statistic [a] | df | P-value (raw) | P-value adj. (Bonferroni)[b] |
|---|---|---|---|---|---|---|---|
| p-tau217 | p-tau217 | 0.188 | 0.171 | 1.098 | 34.252 | 0.280 | 1 |
| p-tau217 | Time × p-tau217 | -0.017 | 0.047 | -0.369 | 71.298 | 0.714 | 1 |
| NfL | NfL | -0.003 | 0.003 | -1.273 | 39.141 | 0.210 | 1 |
| NfL | Time × NfL | -0.001 | 0.002 | -0.473 | 75.919 | 0.638 | 1 |
| GFAP | GFAP | 6.21e-4 | 0.002 | 0.367 | 34.528 | 0.716 | 1 |
| GFAP | Time × GFAP | -7.48e-4 | 5.35e-4 | -1.398 | 71.389 | 0.167 | 0.833 |
| Aβ42/40 | Aβ42/40 | -5.618 | 8.528 | -0.659 | 36.428 | 0.514 | 1 |
| Aβ42/40 | Time × Aβ42/40 | -0.314 | 4.33 | -0.072 | 72.323 | 0.942 | 1 |
| **Visuospatial** | | | | | | | |
| p-tau181 | p-tau181 | -0.007 | 0.01 | -0.657 | 42.781 | 0.515 | 1 |
| p-tau181 | Time × p-tau181 | -0.009 | 0.01 | -0.871 | 82.777 | 0.386 | 1 |
| p-tau217 | p-tau217 | 0.011 | 0.119 | 0.093 | 35.363 | 0.926 | 1 |
| p-tau217 | Time × p-tau217 | 0.054 | 0.049 | 1.114 | 75.063 | 0.269 | 1 |
| NfL | NfL | -0.002 | 0.002 | -0.833 | 46.307 | 0.409 | 1 |
| NfL | Time × NfL | 0.002 | 0.002 | 0.76 | 83.829 | 0.449 | 1 |
| GFAP | GFAP | -5.82e-4 | 0.001 | -0.503 | 35.936 | 0.618 | 1 |
| GFAP | Time × GFAP | 1.31e-4 | 6.31e-4 | 0.208 | 76.079 | 0.836 | 1 |
| Aβ42/40 | Aβ42/40 | 6.27 | 5.917 | 1.06 | 42.256 | 0.295 | 1 |
| Aβ42/40 | Time × Aβ42/40 | 5.175 | 4.802 | 1.078 | 80.442 | 0.284 | 1 |

Time × Biomarker interaction tests whether baseline biomarker levels modify the rate of neuropsychological domain z-score change. ALS = amyotrophic lateral sclerosis; *APOE* = apolipoprotein E; Aβ42/40 = amyloid-β42 to amyloid-β40 ratio; df = degrees of freedom; SE = standard error; GFAP = glial fibrillary acidic protein; NfL = neurofilament light chain; p-tau181 = tau phosphorylated at threonine 181; p-tau217 = tau phosphorylated at threonine 217.

[a] Mixed-effects model fixed-effect t statistic

[b] P-values Bonferroni-adjusted across five biomarkers per domain.

#### ROC AUC results

***Table S10.*** *ROC AUC results for plasma biomarkers and combination ratios across all pairwise diagnostic group comparisons.*

| Biomarker | AUC | 95% CI | n (A/B) | Higher levels |
|---|---|---|---|---|
| **AD vs ALS** | | | | |
| p-tau217/NfL | 0.919 | 0.853–0.972 | 40/40 | AD |
| NfL | 0.892 | 0.817–0.955 | 41/40 | ALS |
| p-tau181/NfL | 0.819 | 0.720–0.906 | 41/40 | AD |
| GFAP | 0.778 | 0.670–0.873 | 41/40 | AD |
| p-tau181/GFAP | 0.755 | 0.640–0.860 | 41/40 | ALS |
| p-tau217 | 0.646 | 0.519–0.760 | 40/40 | AD |
| p-tau217/GFAP | 0.643 | 0.517–0.764 | 40/40 | ALS |
| Aβ42/40 | 0.600 | 0.467–0.728 | 36/39 | ALS |
| p-tau181 | 0.544 | 0.412–0.668 | 41/40 | AD |
| **AD vs CVD-CU** | | | | |
| p-tau217 | 0.929 | 0.871–0.987 | 40/84 | AD |
| p-tau217/NfL | 0.886 | 0.786–0.945 | 40/84 | AD |
| p-tau181 | 0.818 | 0.766–0.904 | 41/85 | AD |
| GFAP | 0.798 | 0.724–0.868 | 41/86 | AD |
| p-tau217/GFAP | 0.756 | 0.676–0.832 | 40/84 | AD |
| p-tau181/NfL | 0.729 | 0.661–0.809 | 41/85 | AD |
| Aβ42/40 | 0.646 | 0.548–0.721 | 36/82 | CVD-CU |
| NfL | 0.626 | 0.539–0.733 | 41/86 | AD |
| p-tau181/GFAP | 0.539 | 0.425–0.604 | 41/85 | AD |
| **AD vs CVD-I** | | | | |
| p-tau217 | 0.806 | 0.712–0.870 | 40/69 | AD |
| p-tau217/NfL | 0.803 | 0.717–0.877 | 40/69 | AD |
| GFAP | 0.762 | 0.686–0.815 | 41/75 | AD |
| p-tau181 | 0.720 | 0.562–0.806 | 41/68 | AD |
| p-tau181/NfL | 0.687 | 0.593–0.771 | 41/68 | AD |
| Aβ42/40 | 0.662 | 0.578–0.761 | 36/71 | CVD-I |
| p-tau217/GFAP | 0.633 | 0.494–0.716 | 40/69 | AD |
| NfL | 0.537 | 0.435–0.623 | 41/75 | AD |
| p-tau181/GFAP | 0.518 | 0.385–0.604 | 41/68 | CVD-I |
| **AD vs FTD** | | | | |
| p-tau217 | 0.787 | 0.687–0.853 | 40/53 | AD |
| p-tau217/NfL | 0.784 | 0.699–0.893 | 40/53 | AD |
| p-tau181/NfL | 0.735 | 0.639–0.815 | 41/53 | AD |
| GFAP | 0.731 | 0.644–0.858 | 41/53 | AD |
| p-tau181 | 0.718 | 0.621–0.795 | 41/53 | AD |

| Biomarker | AUC | 95% CI | n (A/B) | Higher levels |
|---|---|---|---|---|
| p-tau217/GFAP | 0.680 | 0.562–0.763 | 40/53 | AD |
| Aβ42/40 | 0.592 | 0.464–0.709 | 36/49 | FTD |
| p-tau181/GFAP | 0.542 | 0.439–0.656 | 41/53 | AD |
| NfL | 0.526 | 0.436–0.633 | 41/53 | FTD |
| **AD vs HC** | | | | |
| p-tau181 | 0.910 | 0.864–0.954 | 41/44 | AD |
| p-tau217 | 0.903 | 0.828–0.950 | 40/43 | AD |
| p-tau217/NfL | 0.820 | 0.733–0.909 | 40/43 | AD |
| GFAP | 0.815 | 0.747–0.930 | 41/43 | AD |
| NfL | 0.798 | 0.721–0.881 | 41/43 | AD |
| p-tau217/GFAP | 0.777 | 0.666–0.854 | 40/43 | AD |
| p-tau181/NfL | 0.767 | 0.705–0.853 | 41/43 | AD |
| p-tau181/GFAP | 0.672 | 0.578–0.760 | 41/43 | AD |
| Aβ42/40 | 0.633 | 0.520–0.742 | 36/41 | HC |
| **AD vs MCI** | | | | |
| p-tau217 | 0.762 | 0.668–0.812 | 40/85 | AD |
| GFAP | 0.714 | 0.618–0.797 | 41/85 | AD |
| p-tau217/NfL | 0.705 | 0.600–0.779 | 40/85 | AD |
| p-tau181 | 0.693 | 0.602–0.777 | 41/85 | AD |
| NfL | 0.672 | 0.594–0.746 | 41/85 | AD |
| p-tau217/GFAP | 0.615 | 0.540–0.698 | 40/85 | AD |
| p-tau181/NfL | 0.581 | 0.480–0.673 | 41/85 | AD |
| p-tau181/GFAP | 0.514 | 0.402–0.610 | 41/85 | AD |
| Aβ42/40 | 0.468 | 0.341–0.582 | 36/83 | AD |
| **AD vs PD-CU** | | | | |
| p-tau217 | 0.922 | 0.857–0.977 | 40/48 | AD |
| GFAP | 0.871 | 0.805–0.944 | 41/48 | AD |
| p-tau217/NfL | 0.831 | 0.750–0.895 | 40/48 | AD |
| NfL | 0.815 | 0.700–0.913 | 41/48 | AD |
| p-tau181 | 0.784 | 0.676–0.852 | 41/48 | AD |
| Aβ42/40 | 0.731 | 0.612–0.822 | 36/47 | PD-CU |
| p-tau217/GFAP | 0.673 | 0.583–0.804 | 40/48 | AD |
| p-tau181/GFAP | 0.596 | 0.487–0.721 | 41/48 | PD-CU |
| p-tau181/NfL | 0.559 | 0.434–0.652 | 41/48 | AD |
| **AD vs PD-D** | | | | |
| p-tau217 | 0.832 | 0.742–0.914 | 40/26 | AD |
| p-tau217/NfL | 0.817 | 0.711–0.912 | 40/26 | AD |
| GFAP | 0.757 | 0.641–0.859 | 41/26 | AD |
| p-tau181 | 0.732 | 0.639–0.819 | 41/26 | AD |
| p-tau181/NfL | 0.722 | 0.599–0.813 | 41/26 | AD |

| Biomarker | AUC | 95% CI | n (A/B) | Higher levels |
|---|---|---|---|---|
| p-tau217/GFAP | 0.705 | 0.604–0.807 | 40/26 | AD |
| Aβ42/40 | 0.656 | 0.543–0.781 | 36/25 | PD-D |
| NfL | 0.617 | 0.491–0.772 | 41/26 | AD |
| p-tau181/GFAP | 0.493 | 0.364–0.643 | 41/26 | AD |
| **AD vs PD-MCI** | | | | |
| p-tau217 | 0.930 | 0.894–0.971 | 40/51 | AD |
| p-tau217/NfL | 0.876 | 0.807–0.952 | 40/51 | AD |
| GFAP | 0.839 | 0.740–0.899 | 41/51 | AD |
| p-tau181 | 0.791 | 0.679–0.868 | 41/51 | AD |
| p-tau217/GFAP | 0.783 | 0.659–0.871 | 40/51 | AD |
| NfL | 0.747 | 0.695–0.845 | 41/51 | AD |
| Aβ42/40 | 0.653 | 0.536–0.773 | 36/48 | PD-MCI |
| p-tau181/NfL | 0.626 | 0.493–0.710 | 41/51 | AD |
| p-tau181/GFAP | 0.530 | 0.409–0.677 | 41/51 | AD |
| **ALS vs CVD-CU** | | | | |
| NfL | 0.921 | 0.874–0.963 | 40/86 | ALS |
| p-tau217 | 0.833 | 0.779–0.924 | 40/84 | ALS |
| p-tau217/GFAP | 0.812 | 0.740–0.887 | 40/84 | ALS |
| p-tau181 | 0.802 | 0.689–0.884 | 40/85 | ALS |
| p-tau181/GFAP | 0.773 | 0.692–0.857 | 40/85 | ALS |
| p-tau181/NfL | 0.605 | 0.518–0.680 | 40/85 | CVD-CU |
| p-tau217/NfL | 0.590 | 0.469–0.700 | 40/84 | CVD-CU |
| Aβ42/40 | 0.541 | 0.440–0.627 | 39/82 | CVD-CU |
| GFAP | 0.532 | 0.409–0.627 | 40/86 | CVD-CU |
| **ALS vs CVD-I** | | | | |
| NfL | 0.906 | 0.846–0.948 | 40/75 | ALS |
| p-tau181/GFAP | 0.744 | 0.623–0.820 | 40/68 | ALS |
| p-tau217/GFAP | 0.720 | 0.602–0.812 | 40/69 | ALS |
| p-tau181 | 0.720 | 0.622–0.849 | 40/68 | ALS |
| p-tau217 | 0.695 | 0.616–0.759 | 40/69 | ALS |
| p-tau217/NfL | 0.675 | 0.559–0.756 | 40/69 | CVD-I |
| p-tau181/NfL | 0.654 | 0.560–0.736 | 40/68 | CVD-I |
| GFAP | 0.575 | 0.443–0.675 | 40/75 | CVD-I |
| Aβ42/40 | 0.567 | 0.450–0.664 | 39/71 | CVD-I |
| **ALS vs FTD** | | | | |
| NfL | 0.869 | 0.792–0.921 | 40/53 | ALS |
| p-tau181/GFAP | 0.780 | 0.652–0.867 | 40/53 | ALS |
| p-tau217/GFAP | 0.764 | 0.594–0.875 | 40/53 | ALS |
| p-tau181 | 0.717 | 0.613–0.813 | 40/53 | ALS |
| p-tau217 | 0.686 | 0.562–0.782 | 40/53 | ALS |

| Biomarker | AUC | 95% CI | n (A/B) | Higher levels |
|---|---|---|---|---|
| GFAP | 0.639 | 0.555–0.742 | 40/53 | FTD |
| p-tau217/NfL | 0.608 | 0.503–0.711 | 40/53 | FTD |
| p-tau181/NfL | 0.584 | 0.491–0.707 | 40/53 | FTD |
| Aβ42/40 | 0.495 | 0.374–0.585 | 39/49 | ALS |
| **ALS vs HC** | | | | |
| NfL | 0.990 | 0.973–1.000 | 40/43 | ALS |
| p-tau181 | 0.876 | 0.809–0.948 | 40/44 | ALS |
| p-tau181/GFAP | 0.856 | 0.800–0.923 | 40/43 | ALS |
| p-tau217 | 0.813 | 0.731–0.899 | 40/43 | ALS |
| p-tau217/GFAP | 0.812 | 0.713–0.881 | 40/43 | ALS |
| p-tau217/NfL | 0.752 | 0.632–0.846 | 40/43 | HC |
| p-tau181/NfL | 0.606 | 0.490–0.745 | 40/43 | HC |
| GFAP | 0.576 | 0.447–0.677 | 40/43 | ALS |
| Aβ42/40 | 0.544 | 0.420–0.627 | 39/41 | HC |
| **ALS vs MCI** | | | | |
| NfL | 0.945 | 0.900–0.988 | 40/85 | ALS |
| p-tau217/NfL | 0.800 | 0.729–0.880 | 40/85 | MCI |
| p-tau181/GFAP | 0.761 | 0.700–0.848 | 40/85 | ALS |
| p-tau181/NfL | 0.746 | 0.678–0.810 | 40/85 | MCI |
| p-tau217/GFAP | 0.716 | 0.612–0.815 | 40/85 | ALS |
| p-tau181 | 0.711 | 0.618–0.798 | 40/85 | ALS |
| p-tau217 | 0.639 | 0.555–0.749 | 40/85 | ALS |
| GFAP | 0.615 | 0.479–0.730 | 40/85 | MCI |
| Aβ42/40 | 0.565 | 0.436–0.697 | 39/83 | ALS |
| **ALS vs PD-CU** | | | | |
| NfL | 0.985 | 0.945–0.999 | 40/48 | ALS |
| p-tau217 | 0.842 | 0.745–0.922 | 40/48 | ALS |
| p-tau181 | 0.771 | 0.687–0.867 | 40/48 | ALS |
| p-tau181/NfL | 0.766 | 0.683–0.841 | 40/48 | PD-CU |
| p-tau217/GFAP | 0.755 | 0.663–0.834 | 40/48 | ALS |
| p-tau217/NfL | 0.726 | 0.651–0.834 | 40/48 | PD-CU |
| p-tau181/GFAP | 0.693 | 0.605–0.784 | 40/48 | ALS |
| Aβ42/40 | 0.631 | 0.515–0.742 | 39/47 | PD-CU |
| GFAP | 0.556 | 0.394–0.687 | 40/48 | ALS |
| **ALS vs PD-D** | | | | |
| NfL | 0.938 | 0.882–0.987 | 40/26 | ALS |
| p-tau181/GFAP | 0.783 | 0.670–0.864 | 40/26 | ALS |
| p-tau217/GFAP | 0.777 | 0.678–0.863 | 40/26 | ALS |
| p-tau181 | 0.743 | 0.610–0.870 | 40/26 | ALS |
| p-tau217 | 0.733 | 0.598–0.818 | 40/26 | ALS |

| Biomarker | AUC | 95% CI | n (A/B) | Higher levels |
|---|---|---|---|---|
| p-tau217/NfL | 0.708 | 0.596–0.831 | 40/26 | PD-D |
| p-tau181/NfL | 0.659 | 0.547–0.766 | 40/26 | PD-D |
| GFAP | 0.592 | 0.452–0.734 | 40/26 | PD-D |
| Aβ42/40 | 0.469 | 0.320–0.652 | 39/25 | PD-D |
| **ALS vs PD-MCI** | | | | |
| NfL | 0.981 | 0.962–0.998 | 40/51 | ALS |
| p-tau217 | 0.856 | 0.788–0.922 | 40/51 | ALS |
| p-tau217/GFAP | 0.820 | 0.758–0.901 | 40/51 | ALS |
| p-tau181 | 0.775 | 0.655–0.857 | 40/51 | ALS |
| p-tau181/GFAP | 0.753 | 0.640–0.841 | 40/51 | ALS |
| p-tau181/NfL | 0.735 | 0.595–0.827 | 40/51 | PD-MCI |
| p-tau217/NfL | 0.676 | 0.554–0.775 | 40/51 | PD-MCI |
| Aβ42/40 | 0.542 | 0.416–0.673 | 39/48 | PD-MCI |
| GFAP | 0.528 | 0.414–0.622 | 40/51 | ALS |
| **CVD-CU vs CVD-I** | | | | |
| p-tau217 | 0.637 | 0.575–0.730 | 84/69 | CVD-I |
| p-tau217/GFAP | 0.633 | 0.560–0.713 | 84/69 | CVD-I |
| p-tau181 | 0.597 | 0.529–0.686 | 85/68 | CVD-I |
| p-tau217/NfL | 0.596 | 0.497–0.678 | 84/69 | CVD-I |
| NfL | 0.581 | 0.488–0.660 | 86/75 | CVD-I |
| GFAP | 0.558 | 0.484–0.645 | 86/75 | CVD-I |
| p-tau181/GFAP | 0.555 | 0.469–0.641 | 85/68 | CVD-I |
| p-tau181/NfL | 0.554 | 0.477–0.646 | 85/68 | CVD-CU |
| Aβ42/40 | 0.524 | 0.429–0.614 | 82/71 | CVD-I |
| **CVD-CU vs FTD** | | | | |
| NfL | 0.642 | 0.558–0.733 | 86/53 | FTD |
| p-tau217 | 0.631 | 0.536–0.743 | 84/53 | FTD |
| GFAP | 0.628 | 0.539–0.724 | 86/53 | FTD |
| p-tau217/GFAP | 0.596 | 0.518–0.675 | 84/53 | FTD |
| p-tau181 | 0.594 | 0.493–0.667 | 85/53 | FTD |
| Aβ42/40 | 0.553 | 0.461–0.640 | 82/49 | CVD-CU |
| p-tau181/NfL | 0.520 | 0.373–0.616 | 85/53 | FTD |
| p-tau181/GFAP | 0.507 | 0.421–0.618 | 85/53 | CVD-CU |
| p-tau217/NfL | 0.477 | 0.378–0.566 | 84/53 | FTD |
| **CVD-CU vs HC** | | | | |
| p-tau217/NfL | 0.680 | 0.578–0.755 | 84/43 | HC |
| p-tau181 | 0.638 | 0.556–0.753 | 85/44 | CVD-CU |
| NfL | 0.633 | 0.567–0.700 | 86/43 | CVD-CU |
| p-tau181/GFAP | 0.624 | 0.527–0.703 | 85/43 | CVD-CU |
| GFAP | 0.547 | 0.449–0.638 | 86/43 | CVD-CU |

| Biomarker | AUC | 95% CI | n (A/B) | Higher levels |
| --- | --- | --- | --- | --- |
| p-tau217 | 0.525 | 0.423–0.626 | 84/43 | HC |
| p-tau181/NfL | 0.506 | 0.428–0.574 | 85/43 | CVD-CU |
| Aβ42/40 | 0.501 | 0.424–0.636 | 82/41 | CVD-CU |
| p-tau217/GFAP | 0.489 | 0.400–0.586 | 84/43 | HC |
| **CVD-CU vs MCI** | | | | |
| p-tau217/NfL | 0.735 | 0.671–0.790 | 84/85 | MCI |
| p-tau217 | 0.683 | 0.596–0.756 | 84/85 | MCI |
| p-tau181/NfL | 0.653 | 0.575–0.748 | 85/85 | MCI |
| p-tau217/GFAP | 0.632 | 0.569–0.690 | 84/85 | MCI |
| p-tau181 | 0.622 | 0.508–0.700 | 85/85 | MCI |
| Aβ42/40 | 0.612 | 0.532–0.707 | 82/83 | CVD-CU |
| GFAP | 0.598 | 0.522–0.653 | 86/85 | MCI |
| p-tau181/GFAP | 0.551 | 0.484–0.655 | 85/85 | CVD-CU |
| NfL | 0.538 | 0.454–0.616 | 86/85 | CVD-CU |
| **CVD-CU vs PD-CU** | | | | |
| p-tau181/NfL | 0.676 | 0.597–0.753 | 85/48 | PD-CU |
| NfL | 0.665 | 0.573–0.733 | 86/48 | CVD-CU |
| p-tau217/NfL | 0.645 | 0.550–0.719 | 84/48 | PD-CU |
| p-tau181/GFAP | 0.623 | 0.540–0.699 | 85/48 | PD-CU |
| GFAP | 0.598 | 0.498–0.673 | 86/48 | CVD-CU |
| p-tau217/GFAP | 0.590 | 0.494–0.663 | 84/48 | PD-CU |
| Aβ42/40 | 0.588 | 0.494–0.699 | 82/47 | PD-CU |
| p-tau181 | 0.557 | 0.463–0.634 | 85/48 | PD-CU |
| p-tau217 | 0.524 | 0.451–0.607 | 84/48 | PD-CU |
| **CVD-CU vs PD-D** | | | | |
| p-tau217/NfL | 0.630 | 0.530–0.773 | 84/26 | PD-D |
| p-tau217 | 0.602 | 0.438–0.709 | 84/26 | PD-D |
| GFAP | 0.586 | 0.414–0.717 | 86/26 | PD-D |
| p-tau217/GFAP | 0.579 | 0.412–0.656 | 84/26 | PD-D |
| p-tau181 | 0.578 | 0.467–0.688 | 85/26 | PD-D |
| p-tau181/NfL | 0.546 | 0.409–0.641 | 85/26 | CVD-CU |
| p-tau181/GFAP | 0.524 | 0.386–0.628 | 85/26 | CVD-CU |
| NfL | 0.518 | 0.421–0.637 | 86/26 | CVD-CU |
| Aβ42/40 | 0.516 | 0.398–0.633 | 82/25 | CVD-CU |
| **CVD-CU vs PD-MCI** | | | | |
| p-tau181/NfL | 0.634 | 0.550–0.699 | 85/51 | PD-MCI |
| NfL | 0.586 | 0.478–0.677 | 86/51 | CVD-CU |
| p-tau217/NfL | 0.572 | 0.493–0.649 | 84/51 | PD-MCI |
| p-tau181 | 0.570 | 0.491–0.649 | 85/51 | CVD-CU |
| p-tau181/GFAP | 0.556 | 0.424–0.661 | 85/51 | CVD-CU |

| Biomarker | AUC | 95% CI | n (A/B) | Higher levels |
|---|---|---|---|---|
| p-tau217 | 0.543 | 0.413–0.644 | 84/51 | CVD-CU |
| p-tau217/GFAP | 0.512 | 0.411–0.613 | 84/51 | PD-MCI |
| GFAP | 0.510 | 0.435–0.603 | 86/51 | CVD-CU |
| Aβ42/40 | 0.503 | 0.374–0.589 | 82/48 | CVD-CU |
| **CVD-I vs FTD** | | | | |
| Aβ42/40 | 0.579 | 0.497–0.695 | 71/49 | CVD-I |
| p-tau181/NfL | 0.565 | 0.450–0.677 | 68/53 | FTD |
| p-tau181/GFAP | 0.565 | 0.433–0.644 | 68/53 | CVD-I |
| GFAP | 0.564 | 0.461–0.655 | 75/53 | FTD |
| NfL | 0.563 | 0.469–0.640 | 75/53 | FTD |
| p-tau217/NfL | 0.559 | 0.459–0.672 | 69/53 | CVD-I |
| p-tau217/GFAP | 0.541 | 0.467–0.664 | 69/53 | CVD-I |
| p-tau181 | 0.504 | 0.391–0.615 | 68/53 | FTD |
| p-tau217 | 0.502 | 0.426–0.627 | 69/53 | FTD |
| **CVD-I vs HC** | | | | |
| NfL | 0.740 | 0.672–0.819 | 75/43 | CVD-I |
| p-tau181 | 0.727 | 0.627–0.798 | 68/44 | CVD-I |
| p-tau181/GFAP | 0.684 | 0.566–0.785 | 68/43 | CVD-I |
| p-tau217/GFAP | 0.638 | 0.566–0.728 | 69/43 | CVD-I |
| p-tau217 | 0.623 | 0.537–0.695 | 69/43 | CVD-I |
| p-tau181/NfL | 0.577 | 0.493–0.731 | 68/43 | CVD-I |
| p-tau217/NfL | 0.548 | 0.437–0.671 | 69/43 | CVD-I |
| Aβ42/40 | 0.520 | 0.403–0.596 | 71/41 | CVD-I |
| GFAP | 0.515 | 0.394–0.633 | 75/43 | CVD-I |
| **CVD-I vs MCI** | | | | |
| Aβ42/40 | 0.638 | 0.553–0.706 | 71/83 | CVD-I |
| NfL | 0.630 | 0.579–0.709 | 75/85 | CVD-I |
| p-tau217/NfL | 0.620 | 0.532–0.709 | 69/85 | MCI |
| p-tau181/NfL | 0.608 | 0.521–0.705 | 68/85 | MCI |
| GFAP | 0.545 | 0.478–0.616 | 75/85 | MCI |
| p-tau217 | 0.538 | 0.427–0.606 | 69/85 | MCI |
| p-tau181 | 0.522 | 0.439–0.627 | 68/85 | MCI |
| p-tau217/GFAP | 0.509 | 0.434–0.587 | 69/85 | CVD-I |
| p-tau181/GFAP | 0.498 | 0.419–0.576 | 68/85 | CVD-I |
| **CVD-I vs PD-CU** | | | | |
| NfL | 0.755 | 0.673–0.831 | 75/48 | CVD-I |
| GFAP | 0.664 | 0.581–0.752 | 75/48 | CVD-I |
| p-tau217 | 0.647 | 0.545–0.726 | 69/48 | CVD-I |
| p-tau181/NfL | 0.633 | 0.565–0.729 | 68/48 | PD-CU |
| p-tau181/GFAP | 0.581 | 0.480–0.699 | 68/48 | PD-CU |

| Biomarker | AUC | 95% CI | n (A/B) | Higher levels |
|---|---|---|---|---|
| Aβ42/40 | 0.569 | 0.465–0.662 | 71/47 | PD-CU |
| p-tau181 | 0.551 | 0.432–0.642 | 68/48 | CVD-I |
| p-tau217/GFAP | 0.539 | 0.428–0.610 | 69/48 | CVD-I |
| p-tau217/NfL | 0.518 | 0.392–0.630 | 69/48 | CVD-I |
| **CVD-I vs PD-D** | | | | |
| p-tau217/GFAP | 0.570 | 0.458–0.659 | 69/26 | CVD-I |
| NfL | 0.569 | 0.467–0.696 | 75/26 | CVD-I |
| GFAP | 0.532 | 0.408–0.637 | 75/26 | PD-D |
| p-tau217 | 0.528 | 0.373–0.641 | 69/26 | CVD-I |
| Aβ42/40 | 0.526 | 0.443–0.648 | 71/25 | CVD-I |
| p-tau181/NfL | 0.483 | 0.368–0.605 | 68/26 | CVD-I |
| p-tau181 | 0.478 | 0.368–0.619 | 68/26 | CVD-I |
| p-tau181/GFAP | 0.477 | 0.385–0.607 | 68/26 | CVD-I |
| p-tau217/NfL | 0.474 | 0.375–0.598 | 69/26 | CVD-I |
| **CVD-I vs PD-MCI** | | | | |
| NfL | 0.685 | 0.575–0.756 | 75/51 | CVD-I |
| p-tau217 | 0.681 | 0.585–0.772 | 69/51 | CVD-I |
| p-tau217/GFAP | 0.656 | 0.576–0.759 | 69/51 | CVD-I |
| p-tau181/NfL | 0.583 | 0.486–0.694 | 68/51 | PD-MCI |
| GFAP | 0.581 | 0.514–0.681 | 75/51 | CVD-I |
| p-tau217/NfL | 0.548 | 0.473–0.651 | 69/51 | CVD-I |
| p-tau181 | 0.538 | 0.433–0.649 | 68/51 | CVD-I |
| p-tau181/GFAP | 0.513 | 0.414–0.584 | 68/51 | CVD-I |
| Aβ42/40 | 0.483 | 0.416–0.586 | 71/48 | CVD-I |
| **FTD vs HC** | | | | |
| NfL | 0.797 | 0.706–0.871 | 53/43 | FTD |
| p-tau181 | 0.718 | 0.639–0.812 | 53/44 | FTD |
| p-tau217/NfL | 0.631 | 0.536–0.752 | 53/43 | FTD |
| p-tau181/GFAP | 0.627 | 0.529–0.732 | 53/43 | FTD |
| p-tau217 | 0.619 | 0.490–0.721 | 53/43 | FTD |
| GFAP | 0.605 | 0.503–0.704 | 53/43 | FTD |
| p-tau217/GFAP | 0.597 | 0.508–0.693 | 53/43 | FTD |
| Aβ42/40 | 0.553 | 0.421–0.670 | 49/41 | HC |
| p-tau181/NfL | 0.509 | 0.385–0.610 | 53/43 | FTD |
| **FTD vs MCI** | | | | |
| NfL | 0.687 | 0.602–0.740 | 53/85 | FTD |
| p-tau217/NfL | 0.672 | 0.587–0.763 | 53/85 | MCI |
| p-tau181/NfL | 0.653 | 0.537–0.734 | 53/85 | MCI |
| p-tau181/GFAP | 0.562 | 0.479–0.636 | 53/85 | FTD |
| Aβ42/40 | 0.561 | 0.477–0.630 | 49/83 | FTD |

| Biomarker | AUC | 95% CI | n (A/B) | Higher levels |
|---|---|---|---|---|
| p-tau217/GFAP | 0.546 | 0.460–0.640 | 53/85 | MCI |
| p-tau217 | 0.538 | 0.455–0.633 | 53/85 | MCI |
| p-tau181 | 0.523 | 0.397–0.605 | 53/85 | MCI |
| GFAP | 0.517 | 0.458–0.606 | 53/85 | MCI |
| **FTD vs PD-CU** | | | | |
| NfL | 0.819 | 0.700–0.901 | 53/48 | FTD |
| GFAP | 0.731 | 0.614–0.826 | 53/48 | FTD |
| p-tau181/NfL | 0.678 | 0.550–0.793 | 53/48 | PD-CU |
| Aβ42/40 | 0.645 | 0.548–0.740 | 49/47 | PD-CU |
| p-tau217 | 0.643 | 0.549–0.729 | 53/48 | FTD |
| p-tau181/GFAP | 0.637 | 0.545–0.755 | 53/48 | PD-CU |
| p-tau217/NfL | 0.599 | 0.477–0.705 | 53/48 | FTD |
| p-tau181 | 0.551 | 0.470–0.621 | 53/48 | FTD |
| p-tau217/GFAP | 0.496 | 0.378–0.605 | 53/48 | PD-CU |
| **FTD vs PD-D** | | | | |
| NfL | 0.637 | 0.514–0.730 | 53/26 | FTD |
| p-tau217/NfL | 0.595 | 0.492–0.683 | 53/26 | FTD |
| p-tau181/NfL | 0.560 | 0.463–0.677 | 53/26 | FTD |
| p-tau217 | 0.532 | 0.392–0.647 | 53/26 | FTD |
| GFAP | 0.531 | 0.440–0.680 | 53/26 | FTD |
| p-tau181/GFAP | 0.530 | 0.405–0.731 | 53/26 | FTD |
| p-tau217/GFAP | 0.520 | 0.436–0.640 | 53/26 | FTD |
| p-tau181 | 0.480 | 0.371–0.596 | 53/26 | FTD |
| Aβ42/40 | 0.456 | 0.286–0.560 | 49/25 | PD-D |
| **FTD vs PD-MCI** | | | | |
| NfL | 0.751 | 0.648–0.810 | 53/51 | FTD |
| p-tau217 | 0.676 | 0.562–0.754 | 53/51 | FTD |
| GFAP | 0.649 | 0.563–0.754 | 53/51 | FTD |
| p-tau181/NfL | 0.636 | 0.522–0.733 | 53/51 | FTD |
| p-tau217/GFAP | 0.613 | 0.519–0.707 | 53/51 | FTD |
| p-tau181/GFAP | 0.566 | 0.436–0.673 | 53/51 | PD-MCI |
| Aβ42/40 | 0.561 | 0.450–0.650 | 49/48 | PD-MCI |
| p-tau217/NfL | 0.546 | 0.437–0.661 | 53/51 | FTD |
| p-tau181 | 0.541 | 0.433–0.654 | 53/51 | FTD |
| **HC vs MCI** | | | | |
| p-tau181 | 0.738 | 0.663–0.814 | 44/85 | MCI |
| p-tau181/NfL | 0.685 | 0.610–0.778 | 43/85 | MCI |
| p-tau181/GFAP | 0.677 | 0.574–0.783 | 43/85 | MCI |
| p-tau217 | 0.657 | 0.576–0.736 | 43/85 | MCI |
| p-tau217/GFAP | 0.635 | 0.537–0.739 | 43/85 | MCI |

| Biomarker | AUC | 95% CI | n (A/B) | Higher levels |
|---|---|---|---|---|
| Aβ42/40 | 0.614 | 0.512–0.726 | 41/83 | HC |
| p-tau217/NfL | 0.611 | 0.524–0.683 | 43/85 | MCI |
| NfL | 0.598 | 0.529–0.680 | 43/85 | MCI |
| GFAP | 0.572 | 0.486–0.661 | 43/85 | MCI |
| **HC vs PD-CU** | | | | |
| p-tau181/GFAP | 0.745 | 0.656–0.835 | 43/48 | PD-CU |
| p-tau181/NfL | 0.716 | 0.618–0.787 | 43/48 | PD-CU |
| p-tau181 | 0.695 | 0.564–0.815 | 44/48 | PD-CU |
| GFAP | 0.662 | 0.542–0.723 | 43/48 | HC |
| Aβ42/40 | 0.590 | 0.483–0.701 | 41/47 | PD-CU |
| p-tau217/GFAP | 0.584 | 0.490–0.661 | 43/48 | PD-CU |
| NfL | 0.560 | 0.458–0.675 | 43/48 | HC |
| p-tau217 | 0.541 | 0.417–0.657 | 43/48 | HC |
| p-tau217/NfL | 0.479 | 0.373–0.616 | 43/48 | HC |
| **HC vs PD-D** | | | | |
| p-tau181 | 0.701 | 0.597–0.831 | 44/26 | PD-D |
| NfL | 0.675 | 0.560–0.791 | 43/26 | PD-D |
| p-tau181/GFAP | 0.642 | 0.493–0.754 | 43/26 | PD-D |
| p-tau217 | 0.587 | 0.466–0.706 | 43/26 | PD-D |
| p-tau181/NfL | 0.578 | 0.444–0.663 | 43/26 | PD-D |
| p-tau217/GFAP | 0.574 | 0.407–0.692 | 43/26 | PD-D |
| p-tau217/NfL | 0.567 | 0.398–0.690 | 43/26 | HC |
| GFAP | 0.562 | 0.416–0.706 | 43/26 | PD-D |
| Aβ42/40 | 0.506 | 0.342–0.627 | 41/25 | HC |
| **HC vs PD-MCI** | | | | |
| p-tau181 | 0.714 | 0.609–0.823 | 44/51 | PD-MCI |
| p-tau181/GFAP | 0.692 | 0.586–0.798 | 43/51 | PD-MCI |
| p-tau181/NfL | 0.675 | 0.607–0.788 | 43/51 | PD-MCI |
| p-tau217/NfL | 0.612 | 0.443–0.677 | 43/51 | HC |
| NfL | 0.574 | 0.477–0.686 | 43/51 | PD-MCI |
| p-tau217 | 0.571 | 0.438–0.674 | 43/51 | HC |
| GFAP | 0.567 | 0.483–0.673 | 43/51 | HC |
| p-tau217/GFAP | 0.526 | 0.415–0.640 | 43/51 | HC |
| Aβ42/40 | 0.511 | 0.397–0.625 | 41/48 | HC |
| **MCI vs PD-CU** | | | | |
| Aβ42/40 | 0.712 | 0.617–0.784 | 83/47 | PD-CU |
| GFAP | 0.695 | 0.624–0.800 | 85/48 | MCI |
| p-tau217 | 0.682 | 0.583–0.775 | 85/48 | MCI |
| NfL | 0.639 | 0.573–0.737 | 85/48 | MCI |
| p-tau217/NfL | 0.623 | 0.536–0.726 | 85/48 | MCI |

| Biomarker | AUC | 95% CI | n (A/B) | Higher levels |
|---|---|---|---|---|
| p-tau181/GFAP | 0.585 | 0.482–0.663 | 85/48 | PD-CU |
| p-tau181 | 0.576 | 0.500–0.678 | 85/48 | MCI |
| p-tau217/GFAP | 0.547 | 0.416–0.634 | 85/48 | MCI |
| p-tau181/NfL | 0.519 | 0.426–0.581 | 85/48 | PD-CU |
| **MCI vs PD-D** | | | | |
| p-tau217/NfL | 0.645 | 0.548–0.739 | 85/26 | MCI |
| p-tau181/NfL | 0.630 | 0.517–0.715 | 85/26 | MCI |
| Aβ42/40 | 0.628 | 0.551–0.765 | 83/25 | PD-D |
| p-tau217/GFAP | 0.571 | 0.475–0.683 | 85/26 | MCI |
| p-tau217 | 0.566 | 0.436–0.675 | 85/26 | MCI |
| NfL | 0.563 | 0.446–0.662 | 85/26 | MCI |
| p-tau181 | 0.545 | 0.399–0.656 | 85/26 | MCI |
| p-tau181/GFAP | 0.515 | 0.366–0.652 | 85/26 | MCI |
| GFAP | 0.482 | 0.373–0.597 | 85/26 | MCI |
| **MCI vs PD-MCI** | | | | |
| p-tau217 | 0.704 | 0.617–0.771 | 85/51 | MCI |
| p-tau217/NfL | 0.695 | 0.590–0.780 | 85/51 | MCI |
| p-tau217/GFAP | 0.649 | 0.546–0.732 | 85/51 | MCI |
| Aβ42/40 | 0.633 | 0.555–0.747 | 83/48 | PD-MCI |
| GFAP | 0.618 | 0.549–0.686 | 85/51 | MCI |
| p-tau181 | 0.572 | 0.499–0.641 | 85/51 | MCI |
| p-tau181/NfL | 0.531 | 0.458–0.606 | 85/51 | MCI |
| p-tau181/GFAP | 0.512 | 0.461–0.629 | 85/51 | PD-MCI |
| NfL | 0.457 | 0.368–0.552 | 85/51 | MCI |
| **PD-CU vs PD-D** | | | | |
| NfL | 0.708 | 0.615–0.819 | 48/26 | PD-D |
| GFAP | 0.688 | 0.591–0.791 | 48/26 | PD-D |
| p-tau181/NfL | 0.647 | 0.547–0.804 | 48/26 | PD-CU |
| Aβ42/40 | 0.625 | 0.467–0.742 | 47/25 | PD-CU |
| p-tau217 | 0.619 | 0.454–0.727 | 48/26 | PD-D |
| p-tau181/GFAP | 0.599 | 0.487–0.694 | 48/26 | PD-CU |
| p-tau181 | 0.533 | 0.406–0.690 | 48/26 | PD-D |
| p-tau217/NfL | 0.529 | 0.402–0.678 | 48/26 | PD-CU |
| p-tau217/GFAP | 0.519 | 0.402–0.655 | 48/26 | PD-CU |
| **PD-CU vs PD-MCI** | | | | |
| NfL | 0.635 | 0.522–0.724 | 48/51 | PD-MCI |
| p-tau217/GFAP | 0.605 | 0.524–0.728 | 48/51 | PD-CU |
| GFAP | 0.604 | 0.495–0.720 | 48/51 | PD-MCI |
| Aβ42/40 | 0.600 | 0.500–0.661 | 47/48 | PD-CU |
| p-tau217/NfL | 0.585 | 0.452–0.699 | 48/51 | PD-CU |

| Biomarker | AUC | 95% CI | n (A/B) | Higher levels |
|---|---|---|---|---|
| p-tau181/GFAP | 0.571 | 0.499–0.671 | 48/51 | PD-CU |
| p-tau181/NfL | 0.552 | 0.438–0.646 | 48/51 | PD-CU |
| p-tau217 | 0.531 | 0.411–0.639 | 48/51 | PD-CU |
| p-tau181 | 0.512 | 0.421–0.628 | 48/51 | PD-CU |
| **PD-D vs PD-MCI** | | | | |
| p-tau217 | 0.652 | 0.524–0.743 | 26/51 | PD-D |
| NfL | 0.618 | 0.485–0.791 | 26/51 | PD-D |
| GFAP | 0.615 | 0.465–0.738 | 26/51 | PD-D |
| p-tau181/NfL | 0.609 | 0.440–0.745 | 26/51 | PD-MCI |
| p-tau217/GFAP | 0.591 | 0.445–0.722 | 26/51 | PD-D |
| p-tau217/NfL | 0.558 | 0.449–0.658 | 26/51 | PD-D |
| p-tau181/GFAP | 0.524 | 0.404–0.641 | 26/51 | PD-MCI |
| p-tau181 | 0.522 | 0.411–0.637 | 26/51 | PD-D |
| Aβ42/40 | 0.504 | 0.366–0.623 | 25/48 | PD-D |

AUC > 0.80 is the descriptive threshold for inclusion in Figure 3. Higher levels = group with the higher mean biomarker value; n (A/B) = participants in the first and second comparison groups, respectively. AUC = area under the receiver operating characteristic curve; Aβ42/40 = amyloid-β42 to amyloid-β40 ratio; CI = confidence interval; GFAP = glial fibrillary acidic protein; NfL = neurofilament light chain; p-tau181 = tau phosphorylated at threonine 181; p-tau217 = tau phosphorylated at threonine 217; AD = Alzheimer's disease; ALS = amyotrophic lateral sclerosis; CVD-CU = cerebrovascular disease, cognitively unimpaired; CVD-I = cerebrovascular disease, cognitively impaired; FTD = frontotemporal dementia; HC = healthy controls; MCI = mild cognitive impairment; PD-CU = Parkinson's disease, cognitively unimpaired; PD-D = Parkinson's disease with dementia; PD-MCI = Parkinson's disease with mild cognitive impairment.

### Supplementary Figures

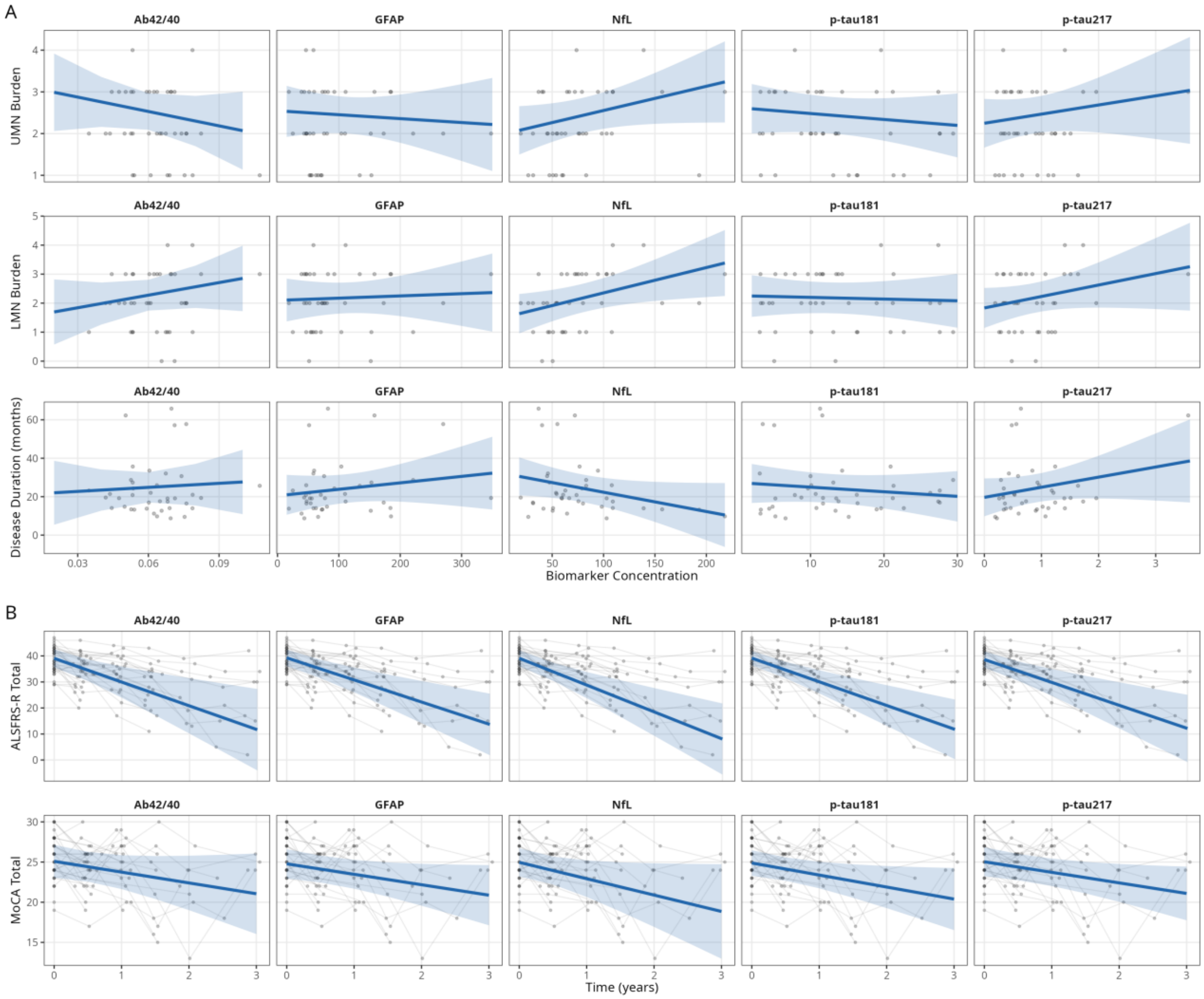


***Supplementary Figure 1. Biomarker-clinical associations. (A)*** *Baseline model associations between plasma biomarkers and clinical outcomes (UMN burden, LMN burden, disease duration).* ***(B)*** *Longitudinal model associations between baseline plasma biomarkers and clinical outcomes (ALSFRS-R decline, MoCA change).*

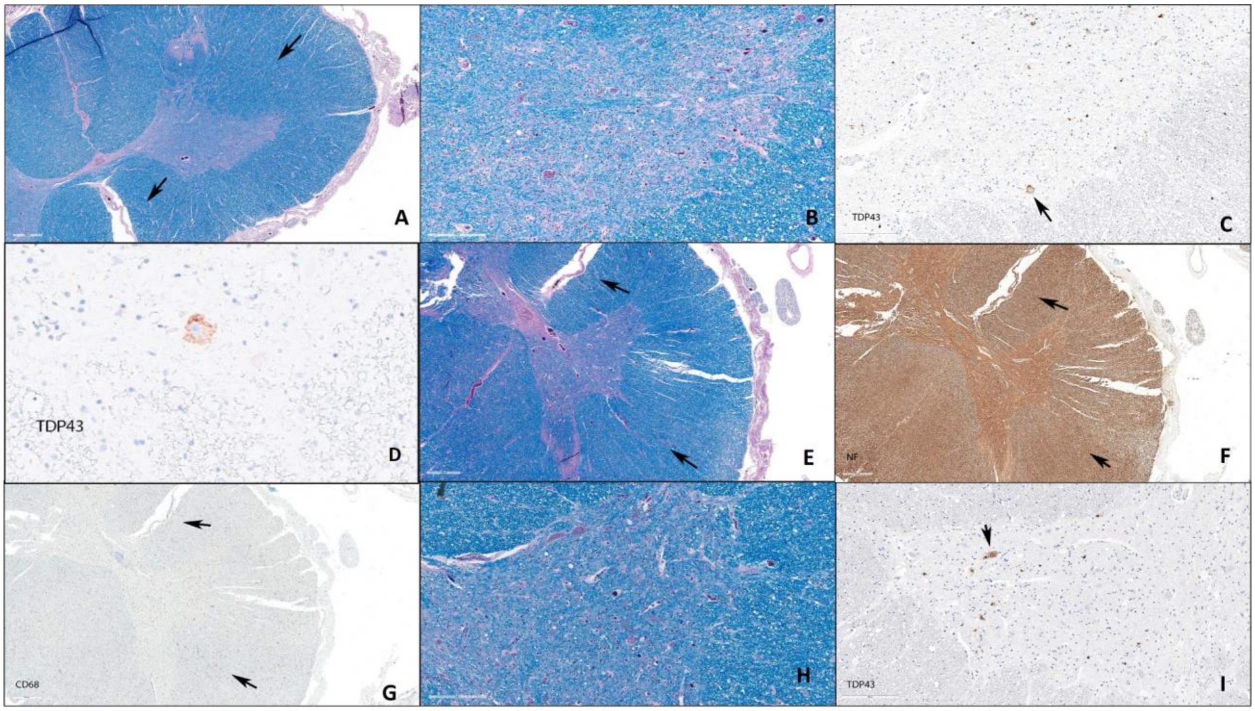


***Supplementary Figure 2. Neuropathology of the spinal cord. (A)*** *H&E/LFB stained slide of the cervical spinal cord showing no corticospinal tract degeneration (arrows);* ***(B)*** *Severe neuronal depletion of the cervical anterior horns;* ***(C,D)*** *remaining cervical motor neurons contain TDP43 immunopositive filamentous cytoplasmic inclusions. Lower thoracic/upper lumbar spinal cord showing no corticospinal tract degeneration on H&E/LFB* ***(E)****, NF* ***(F)*** *or CD68* ***(G)*** *immunolabeling (arrows).* ***(H)*** *Severe neuronal depletion of the anterior horns in the lumbar spinal cord with remaining neurons containing filamentous TDP43 immunopositive cytoplasmic inclusions* ***(I)****.*

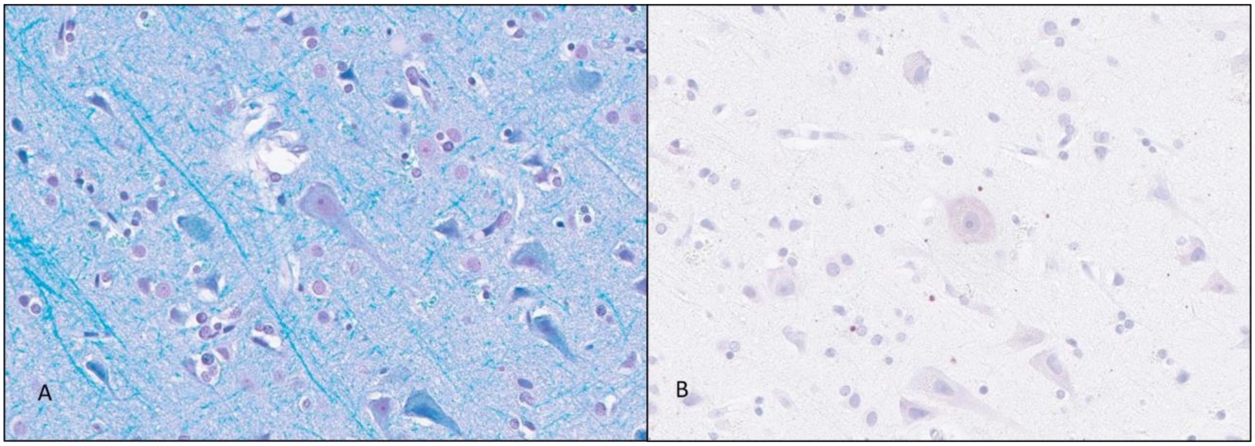


***Supplementary Figure 3. Neuropathology of the motor cortex. (A)*** *On routine staining there was a normal density of Betz neurons within the primary motor cortex and no evidence of atrophy or gliosis;* ***(B)*** *The Betz neurons did not contain TDP43 immunopositive inclusions.*

### ALS Neuropathology Case

A 71-year-old retired civil servant presented to the neuromuscular clinic with a two-year history of insidious onset asymmetrical right upper limb weakness with accompanying loss of muscle bulk, which progressed to involve the left upper extremity. By the time of his first review there was already significant functional compromise of both manual dexterity and upper extremity proximal strength.

Medical background was broadly unremarkable, with a history of treated hypercholesterolemia and benign prostatic hypertrophy only. There was no associated radicular discomfort, dysphagia, dysarthria nor reported cognitive or behavioural change. Mobility was unrestricted.

On examination, there were no bulbar signs. He had atrophy of the musculature of the shoulder girdle (particularly supraspinatus and infraspinatus) and of both first dorsal interossei. Fasciculations were seen in the arms and forearms. He had global weakness in the arms, but lower limb power was retained. Deep Tendon Reflexes were diminished in the arms but enhanced in the lower limbs although there were flexor plantar responses. Sensory examination was normal. A MoCA in clinic was initially scored as 22/30 but once done as part of the ONDRI study was scored as 26/30. While the initial MoCA score was 22/30, there were no subjective cognitive or behavioural complaints nor did the research neuropsychological testing show any objective deficits. He did not meet clinical criteria for Mild Cognitive or Mild Behavioural Impairment, nor frontotemporal dementia/ amyotrophic lateral sclerosis (ALS).

Standard laboratory testing was non-contributory. MRI of the brain was normal. MRI of the spinal cord demonstrated multi-level degenerative changes in the cervical spine but without significant compromise of the central canal or neural foramina. Electromyography studies showed diffuse denervation in both the upper and lower limbs. A clinical diagnosis of limb-onset amyotrophic lateral sclerosis was made, and he was subsequently commenced on Riluzole and went on to receive multidisciplinary care. His Amyotrophic Lateral Sclerosis Functional Rating Scale was scored as 35. P-tau217, p-Tau181, NfL and GFAP were measured at 0.93, 13.1, 47.4 and 153 pg/mL, respectively. *APOE* genotype was ε4/ε3.

Four years after symptom onset, when profound disability had accrued to the extent he no longer had any functional use of the upper limbs and was fully dependent for all activities of daily living, he was referred to the palliative care team for further supportive management. Ultimately, he chose to pursue medical assistance in dying and came to autopsy.

The final neuropathologic diagnosis was Amyotrophic Lateral Sclerosis with predominantly lower motor neuron degeneration and TDP43 immunopathology with low level Alzheimer's type neuropathologic change.

As per the patient's expressed consent, autopsy limited to the brain and spinal cord was performed, and the neuropathology is shown in Supplementary Figures 2 and 3. Upon the patient's death their brain and spinal cord were removed, the right cerebral hemisphere, right hemi-brainstem and cerebellum, and a segment of cervical, thoracic and lumbar spinal cord were frozen, and the remainder of the brain and spinal cord were fixed in formalin. After formalin fixation for greater than two weeks dissection of the left cerebral hemisphere, left hemibrainstem and cerebellum, and remaining spinal cord was performed by an experienced neuropathologist (JK) following a standard neurodegenerative blocking and staining method (the ADNI dementia protocol plus extensive sampling of the brainstem and cervical, thoracic and lumbar spinal cord stained with H&E/LFB, TDP43, p62, tau (AT8), beta-amyloid, NF and CD68 immunohistochemistry).

On macroscopic examination there was mild cortical gyral atrophy and the anterior spinal nerve roots were thin and grey in appearance compared with the normal posterior spinal nerve roots. Microscopic examination of the spinal cord revealed severe neuronal depletion of the anterior horns (Supplementary Figure 2B, H) with remaining alpha-motor neurons containing Bunina bodies and filamentous neuronal cytoplasmic inclusions that label with both TDP43 and p62 (Supplementary Figure 2C, D, I). The anterior spinal nerve roots were thin and pale on myelin staining. There was no evidence of corticospinal tract degeneration seen in the spinal cord on routine staining or immunolabeling with NF or CD68 (Supplementary Figure 2A, E, F, G). Brainstem sections showed the hypoglossal nucleus to have a normal neuronal density with some neurons containing filamentous cytoplasmic inclusions that express TDP43 and p62 and the medullary pyramid was not degenerated.

The cerebral cortex was relatively normal on routine staining including the primary motor cortex in which a normal density of Betz neurons was seen which did not contain TDP43 immunopositive inclusions (Supplementary Figure 3A, B). There was no evidence of TDP43 immunopathology in the mesial temporal structures, nor in the frontal or temporal cortices. Mild Alzheimer's type neuropathologic change (A1B1C1) was present[1]. The internal capsule was not degenerated.

The final neuropathological diagnosis was Amyotrophic Lateral Sclerosis, predominantly lower motor neuron degeneration with TDP43 immunopositive inclusions and Alzheimer's type neuropathologic change (ADNC), A1B1C1.

In our case with a confirmed tissue diagnosis, who had clinically presented with limb onset ALS, and specifically with unimpaired cognition, both p-tau species were found to be demonstrably high in plasma. NfL & GFAP levels were also found to be elevated. Histopathological examination of the spinal cord demonstrated severe loss of motor neurons in the lumbar anterior horns (with remaining cells harbouring neuronal cytoplasmic inclusions containing TDP43) and with thinning of the anterior spinal nerve roots. Corticospinal tract integrity was retained. Interestingly on brain examination, there was no evidence of FTLD-TDP However, Alzheimer's Disease neuropathological change (ADNC) classified as A1B1C1 was observed in the case. This is considered a "low" level, whereas only "intermediate" or "high" range ADNC would be regarded sufficient to account for manifest cognitive impairment[2]. This is consistent with the belief that it is NFT accumulation, and not amyloid plaque burden, that most closely correlates with clinical severity[1].
